# Viral cultures for COVID-19 infectivity assessment – a systematic review (Update 4)

**DOI:** 10.1101/2020.08.04.20167932

**Authors:** T Jefferson, EA Spencer, J Brassey, C Heneghan

## Abstract

**Objective:** to review the evidence from studies comparing SARS-CoV-2 culture, the best indicator of current infection and infectiousness with the results of reverse transcriptase polymerase chain reaction (RT-PCR).

**Methods:** We searched LitCovid, medRxiv, Google Scholar and the WHO Covid-19 database for Covid-19 using the terms ‘viral culture’ or ‘viral replication’ and associated synonyms up to 10 September 2020. We carried out citation matching and included studies reporting attempts to culture or observe SARS-CoV-2 matching with cutoffs for RT-PCR positivity. One reviewer extracted data for each study and a second reviewer checked end edited the extraction and summarised the narratively by sample: fecal, respiratory, environment or mixed.

Where necessary we wrote to corresponding authors of the included or background papers for additional information. We assessed quality using a modified QUADAS 2 risk of bias tool.

This review is part of an <u>Open Evidence Review</u> on Transmission Dynamics of COVID-19. Summaries of the included studies and the protocol (v1) are available at: https://www.cebm.net/evidence-synthesis/transmission-dynamics-of-covid-19/. Searches are updated every 2 weeks. This is the fourth version of this review that was first published on the 4th of August and updated on the 21t of August

**Results:** We included 29 studies reporting culturing or observing tissue invasion by SARS-CoV in sputum, naso or oropharyngeal, urine, stool, blood and environmental samples from patients diagnosed with Covid-19. The data are suggestive of a relation between the time from collection of a specimen to test, cycle threshold and symptom severity. The quality of the studies was moderate with lack of standardised reporting.

Twelve studies reported that Ct values were significantly lower and log copies higher in samples producing live virus culture. Five studies reported no growth in samples based on a Ct cut-off value. These values ranged from CT > 24 for no growth to Ct ≥ 34. Two studies report a strong relationship between Ct value and ability to recover infectious virus and that the odds of live virus culture reduced by 33% for every one unit increase in Ct. A cut-off RT-PCR Ct > 30 was associated with non-infectious samples. One study that analysed the NSP, N and E gene fragments of the PCR result reported different cut-off thresholds depending on the gene fragment analysed. The duration of RNA shedding detected by PCR was far longer compared to detection of live culture. Six out of eight studies reported RNA shedding for longer than 14 days. Yet, infectivity declines after day 8 even among cases with ongoing high viral loads. A very small proportion of people re-testing positive after hospital discharge or with high Ct are likely to be infectious.

**Conclusion:** Prospective routine testing of reference and culture specimens are necessary for each country involved in the pandemic to establish the usefulness and reliability of PCR for Covid-19 and its relation to patients’ factors. Infectivity is related to the date of onset of symptoms and cycle threshold level.

A binary Yes/No approach to the interpretation RT-PCR unvalidated against viral culture will result in false positives with possible segregation of large numbers of people who are no longer infectious and hence not a threat to public health.

## Introduction

The ability to make decisions on the prevention and management of COVID-19 infections rests on our capacity to identify those who are infected and infectious. In the absence of predictive clinical signs or symptoms^1^, the most widely used means of detection is molecular testing using Reverse Transcriptase quantitative Polymerase Chain Reaction (RT-qPCR)^2 3^.

The test amplifies genomic sequences identified in samples. As it is capable of generating observable signals from small samples, it is very sensitive. Amplification of genomic sequence is measured in cycle thresholds (Ct). There appears to be a correlation between Ct values from respiratory samples, symptom onset to test (STT) date and positive viral culture. The lower the Ct value and the shorter the STT, the higher the infectivity potential^4^.

Whether probing for sequences or whole genomes^5^, in the diagnosis of Covid-19 a positive RT-qPCR cannot tell you whether the person is infectious or when the infection began, nor the provenance of the genetic material. Very early in the COVID-19 outbreak it was recognised that cycle threshold values may be a proxy for quantitative measure of viral load, but correlation with clinical progress and transmissibility was not yet known^6^. A positive result indicates that a person has come into contact with the genomic sequence or some other viral antigen at some time in the past. However, presence of viral genome on its own is not sufficient proof of infectivity and caution is needed when evaluating the infectivity of specimens simply based on the detection of viral nucleic acids^5^. In addition, viral genomic material can be still be present weeks after infectious viral clearance.^7^ Like all tests, RT-qPCR requires validation against a gold standard. In this case isolation of a whole virion (as opposed to fragments) and proof that the isolate is capable of replicating its progeny in culture cells is the closest we are likely to get to a gold standard.^8^ The inability of PCR to distinguish between the shedding of live virus or of viral debris, means that is cannot measure a person’s viral load (or quantity of virus present in a person’s excreta).

Our <u>Open Evidence Review</u> of transmission modalities of SARS CoV-2 identified a low number of studies which have attempted viral culture. There are objective difficulties in doing such cultures such as the requirement for a level III laboratory, avoidance of contamination, time and the quality of the specimens as well as financial availability of reagents and culture media to rule out the presence of other pathogens.

As viral culture represents the best indicator of infection and infectiousness, we set out to review the evidence on viral culture compared to PCR, and report the results of those studies attempting viral culture regardless of source (specimen type) of the sample tested.

## Methods

We searched four main databases: LitCovid, medRxiv, Google Scholar and the WHO Covid-19 database for Covid-19 using the terms ‘viral culture’ or ‘viral replication’ and associated synonyms. Searches were last updated on 10 September 2020. Searches are conducted on a per calendar month basis and for databases which do not support such date granularity, the date of publication is approximated. For articles that looked particularly relevant, citation matching was undertaken and relevant results were identified.

We included studies reporting attempts to culture SARS-CoV-2 and those which also estimated the infectiousness of the isolates or observed tissue invasion by SARS CoV-2. One reviewer extracted data for each study and a second review checked end edited the extraction. We tabulated the data and summarised data narratively by mode of sample: fecal, respiratory, environment or mixed.

Where necessary we wrote to corresponding authors of the included or background papers for additional information. We assessed quality using a modified QUADAS 2 risk of bias tool. We simplified the tool as the included studies were not designed as primary diagnostic accuracy studies.^9^

This review is part of an Open Evidence Review on Transmission Dynamics of COVID-19. Summaries of the included studies and the protocol (v1) are available at: https://www.cebm.net/evidence-synthesis/transmission-dynamics-of-covid-19/. Searches are updated every 2 weeks.

This is the fourth update of this review with the addition of four studies identified in the two weeks since the last update.

## Results

We identified 145 articles of possible interest and after screening full texts included 29 (see PRISMA^10^ flow chart - Figure 1). We identified one unpublished study which was not included as no permission to do so was given by the authors. The salient characteristics of each included study are shown in Table 1.

**Figure 1 -.**
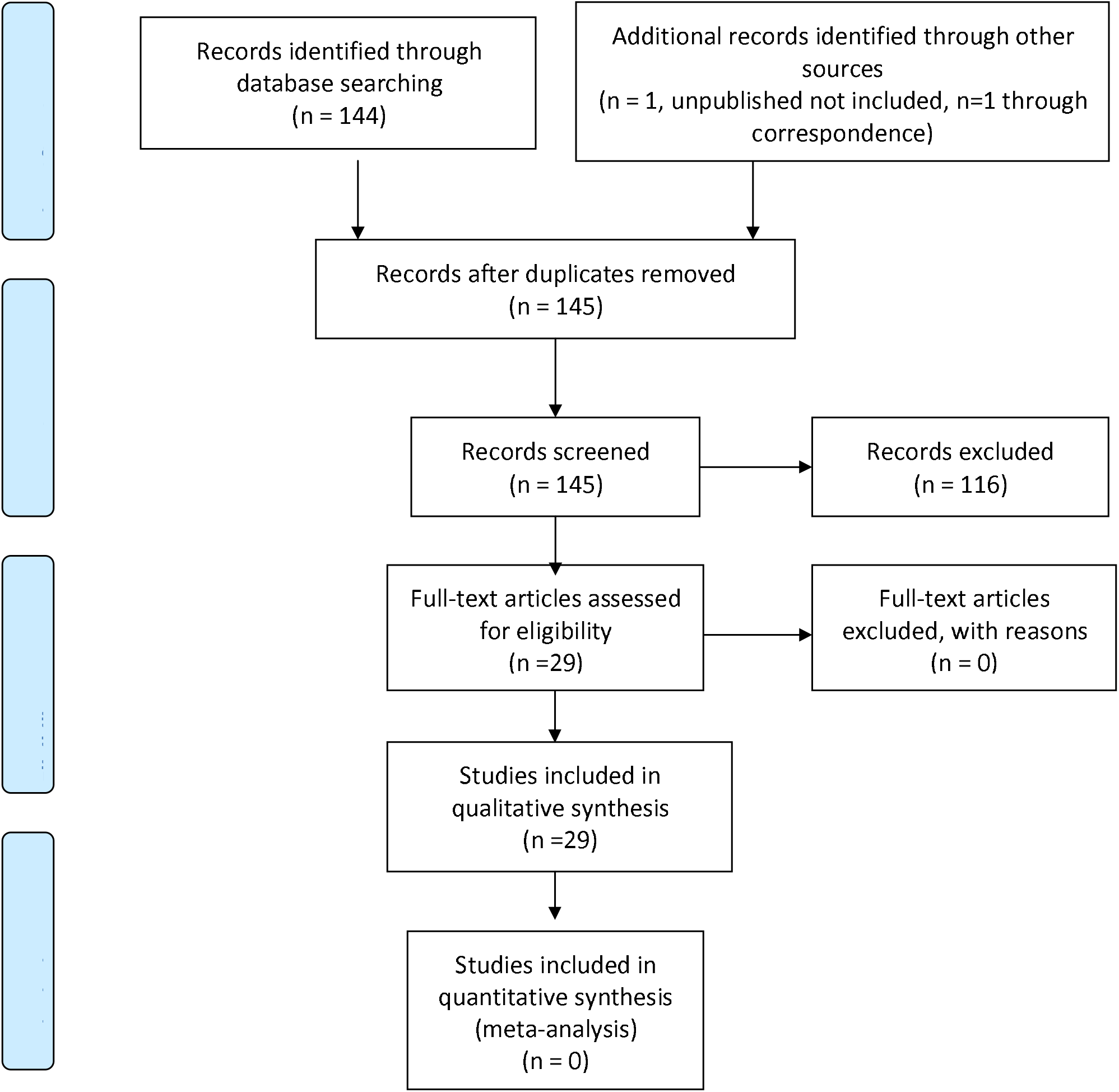
PRISMA 2009 Flow Diagram.

**Table 1.**
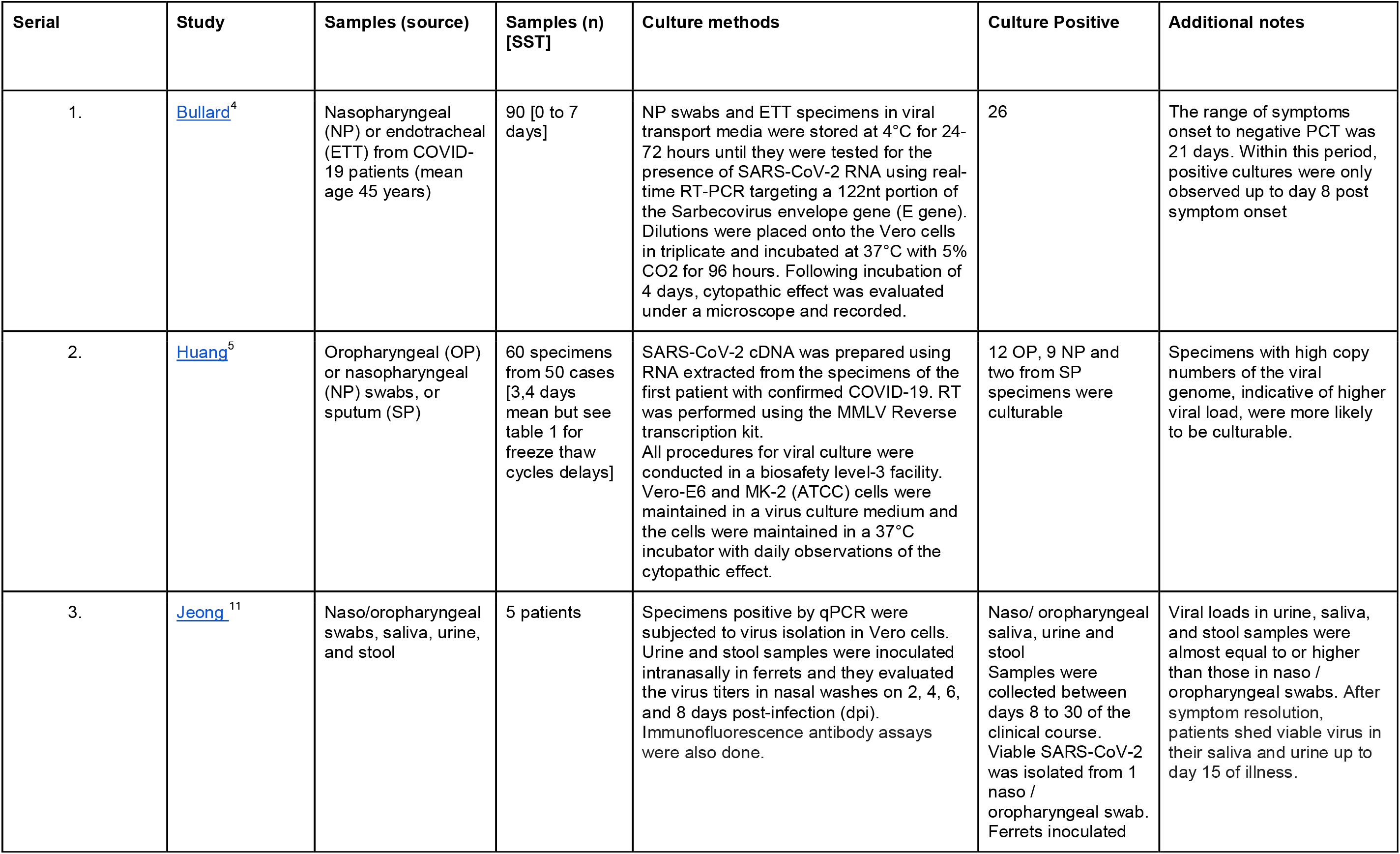
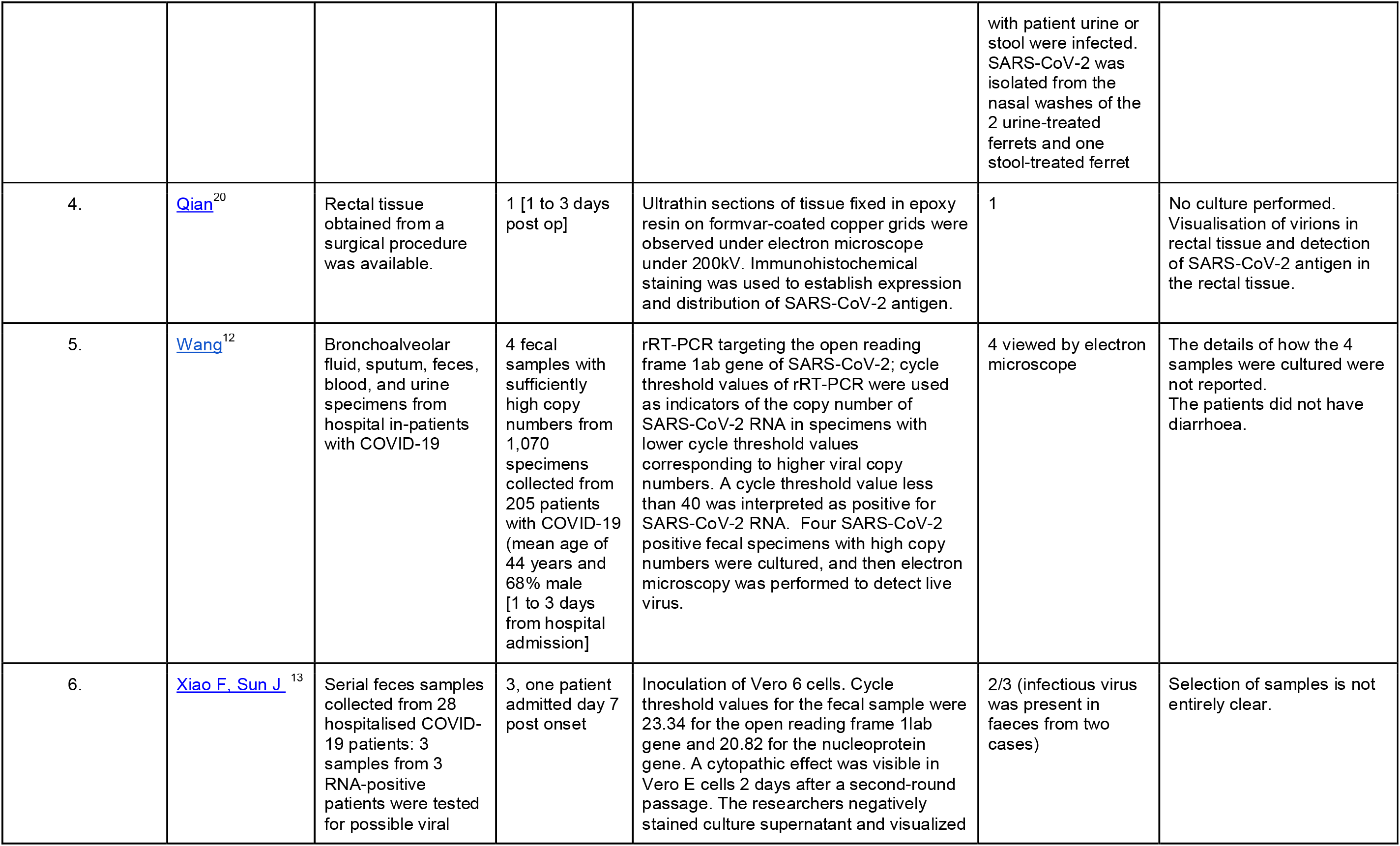
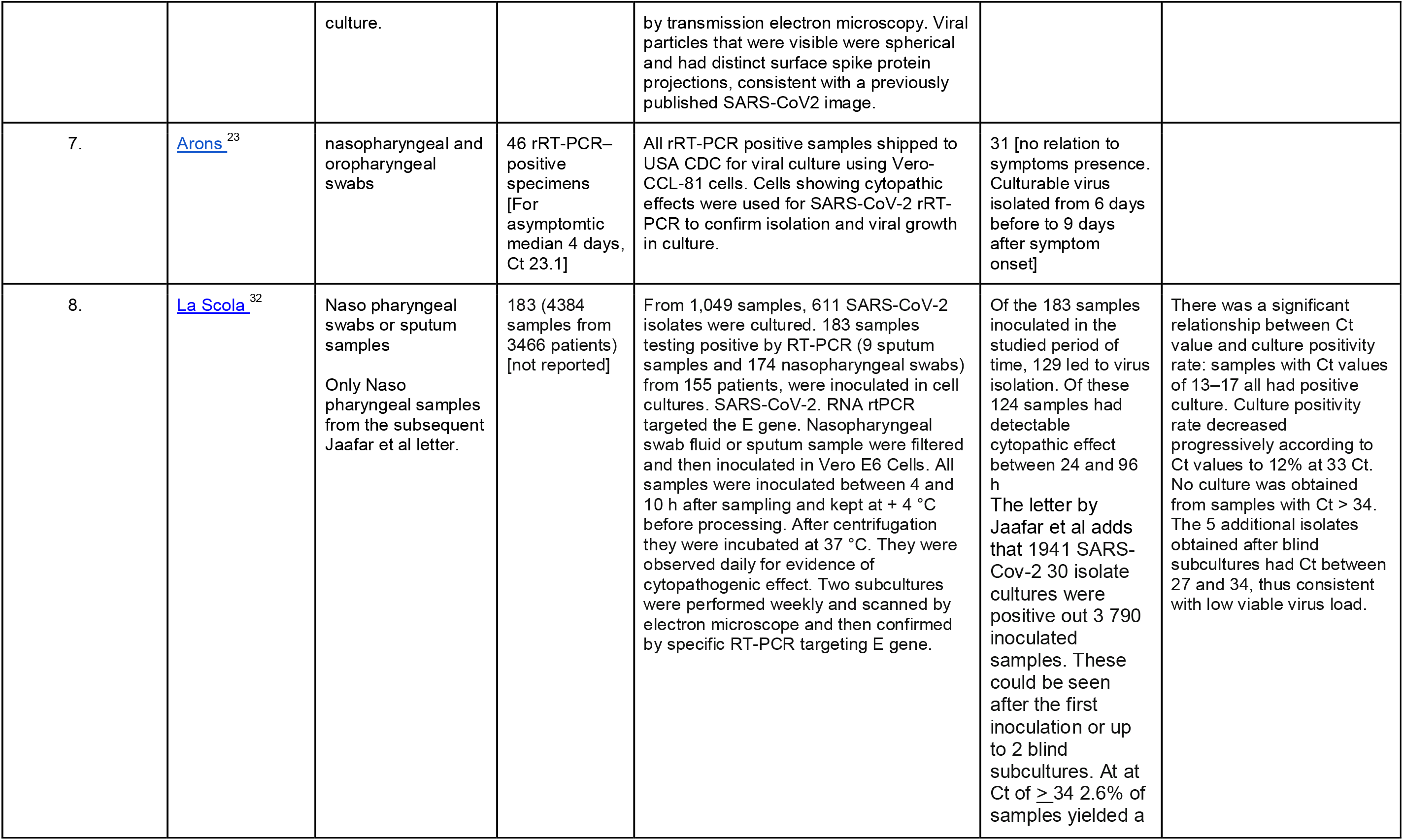
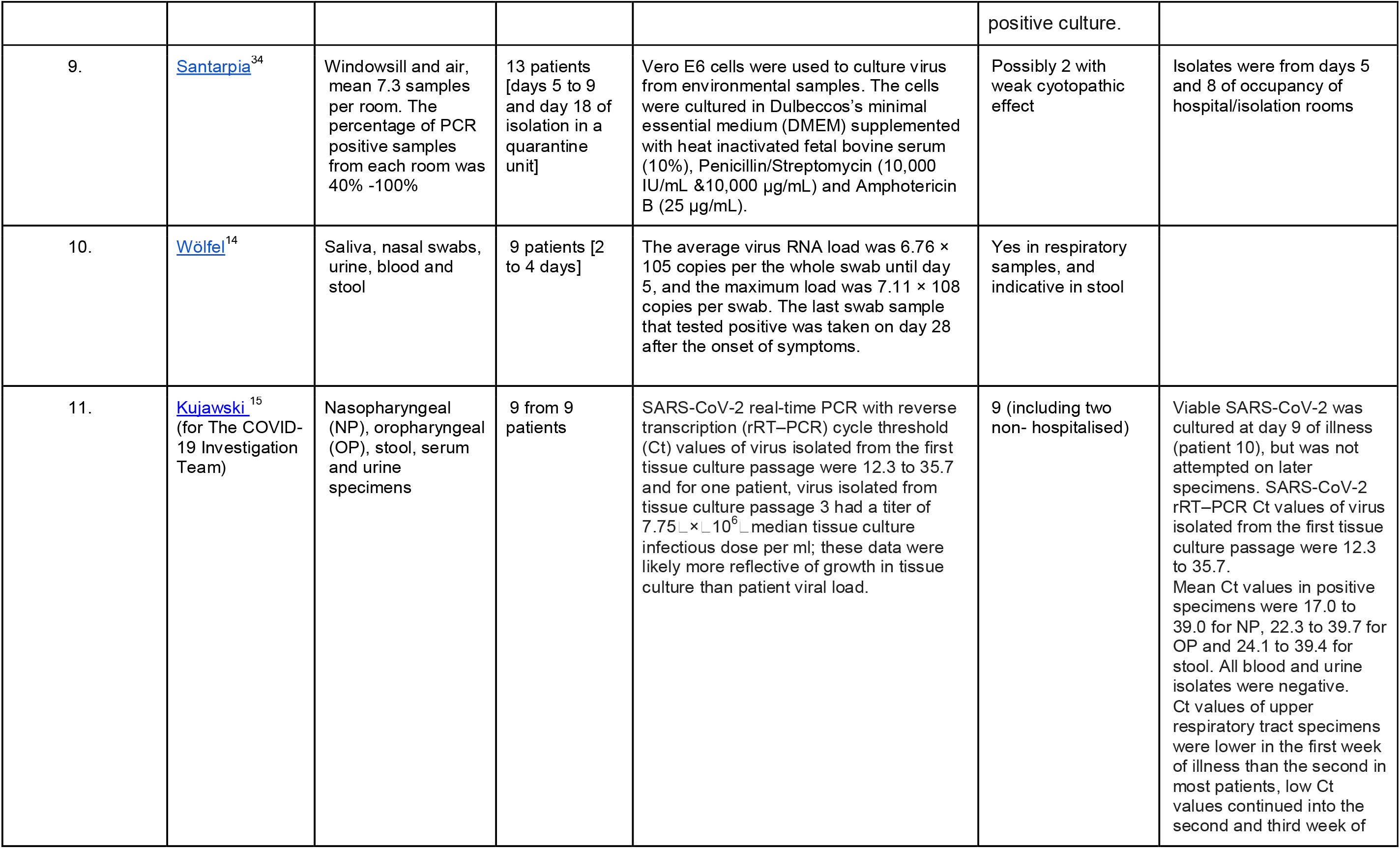
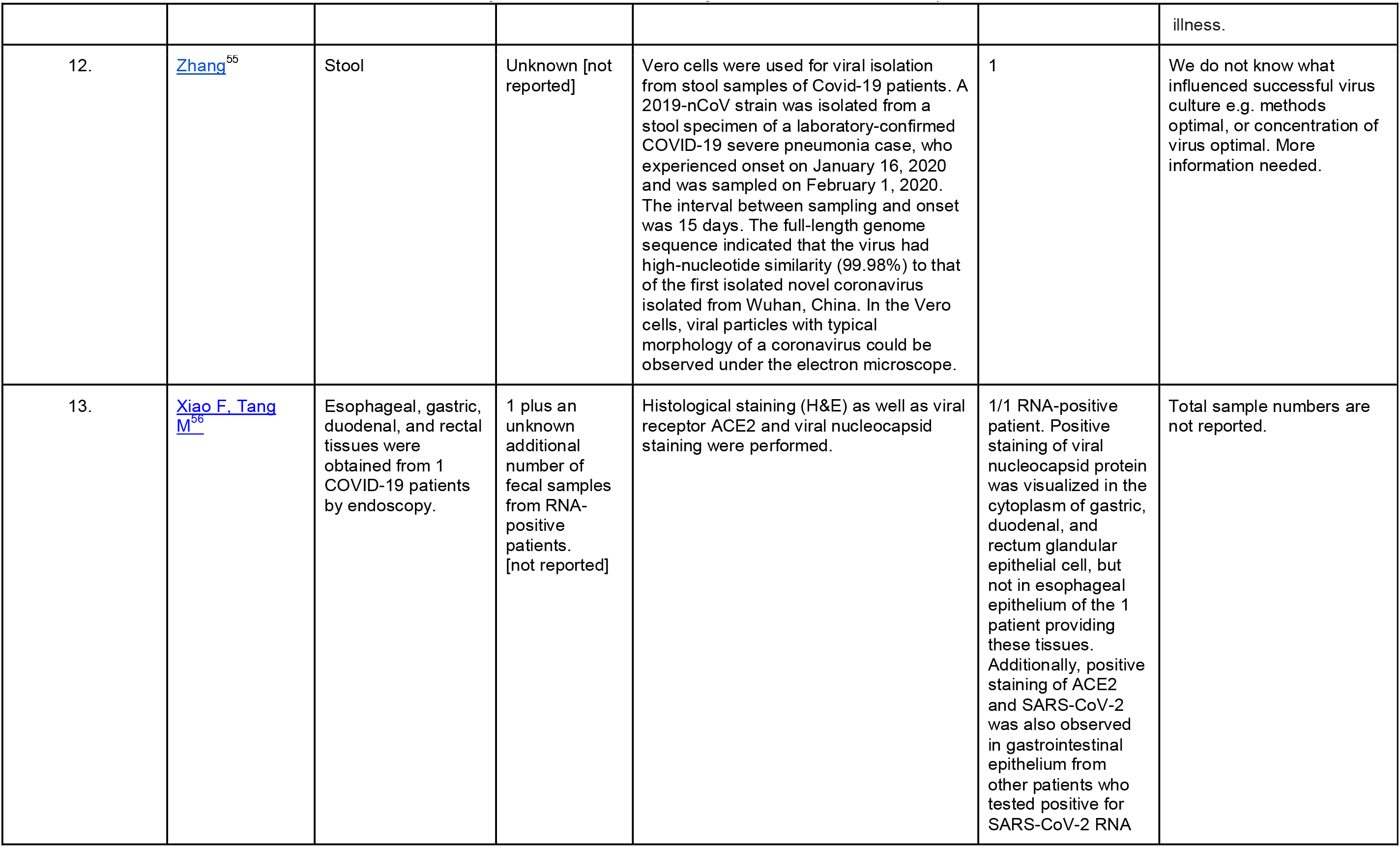
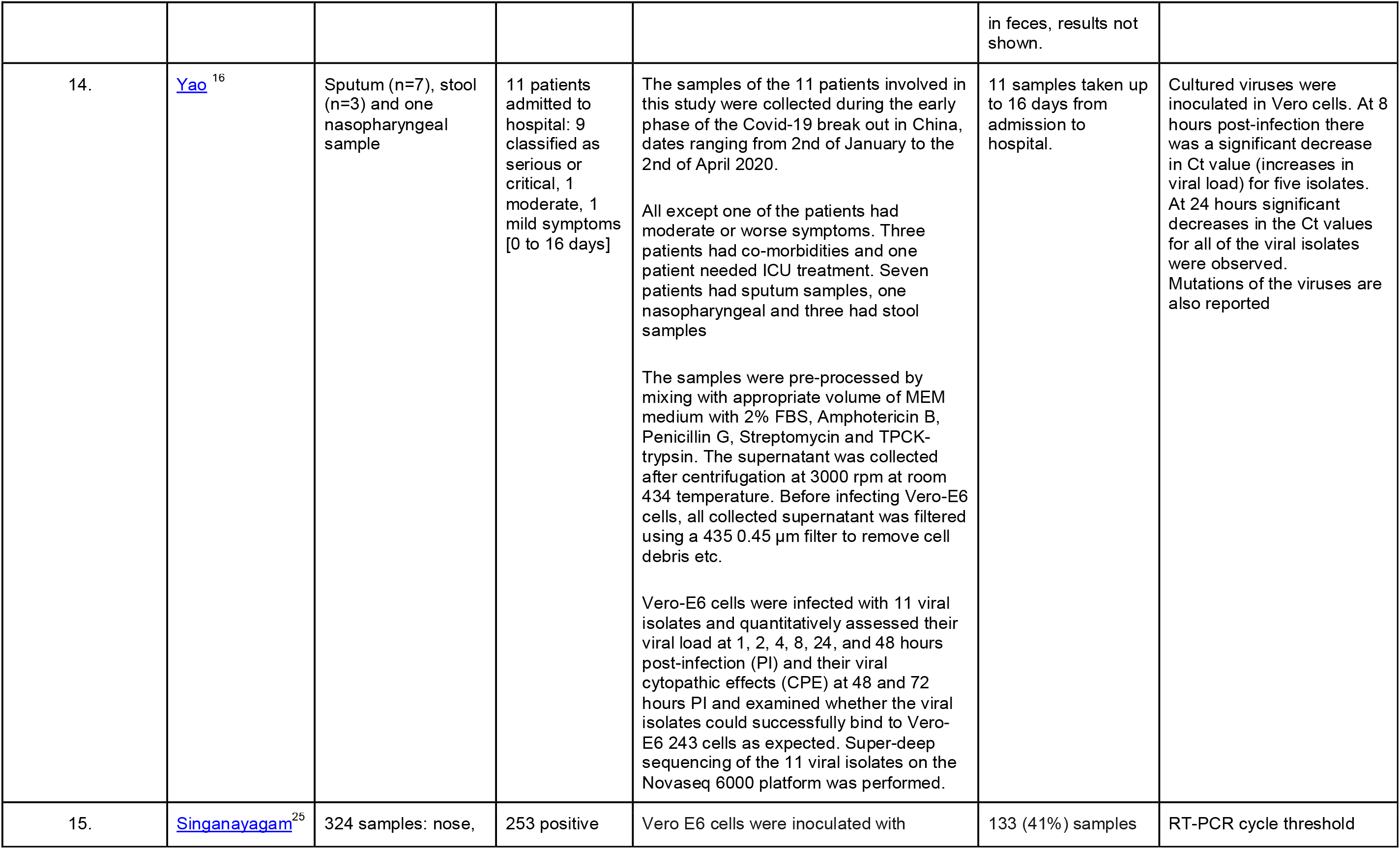
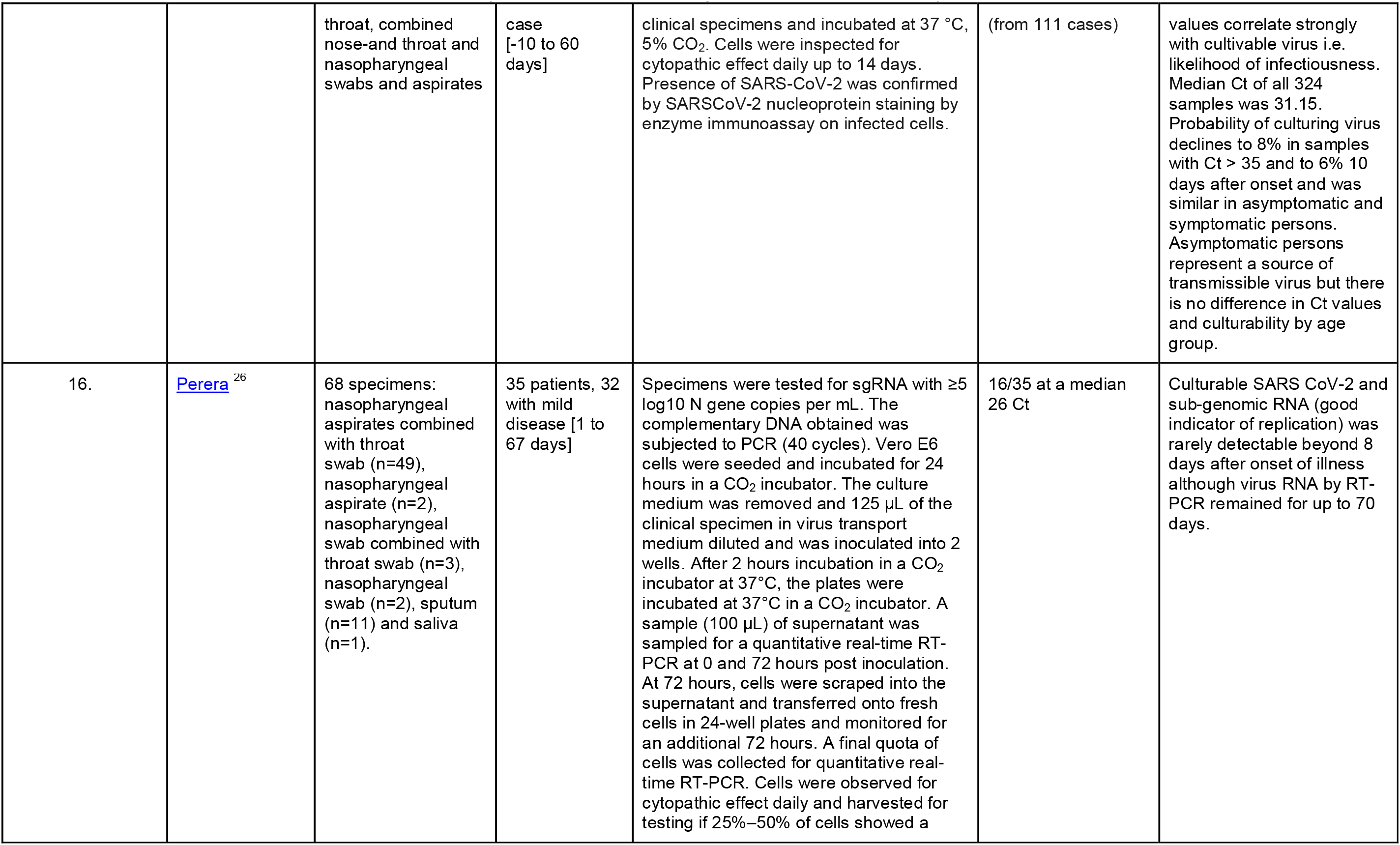
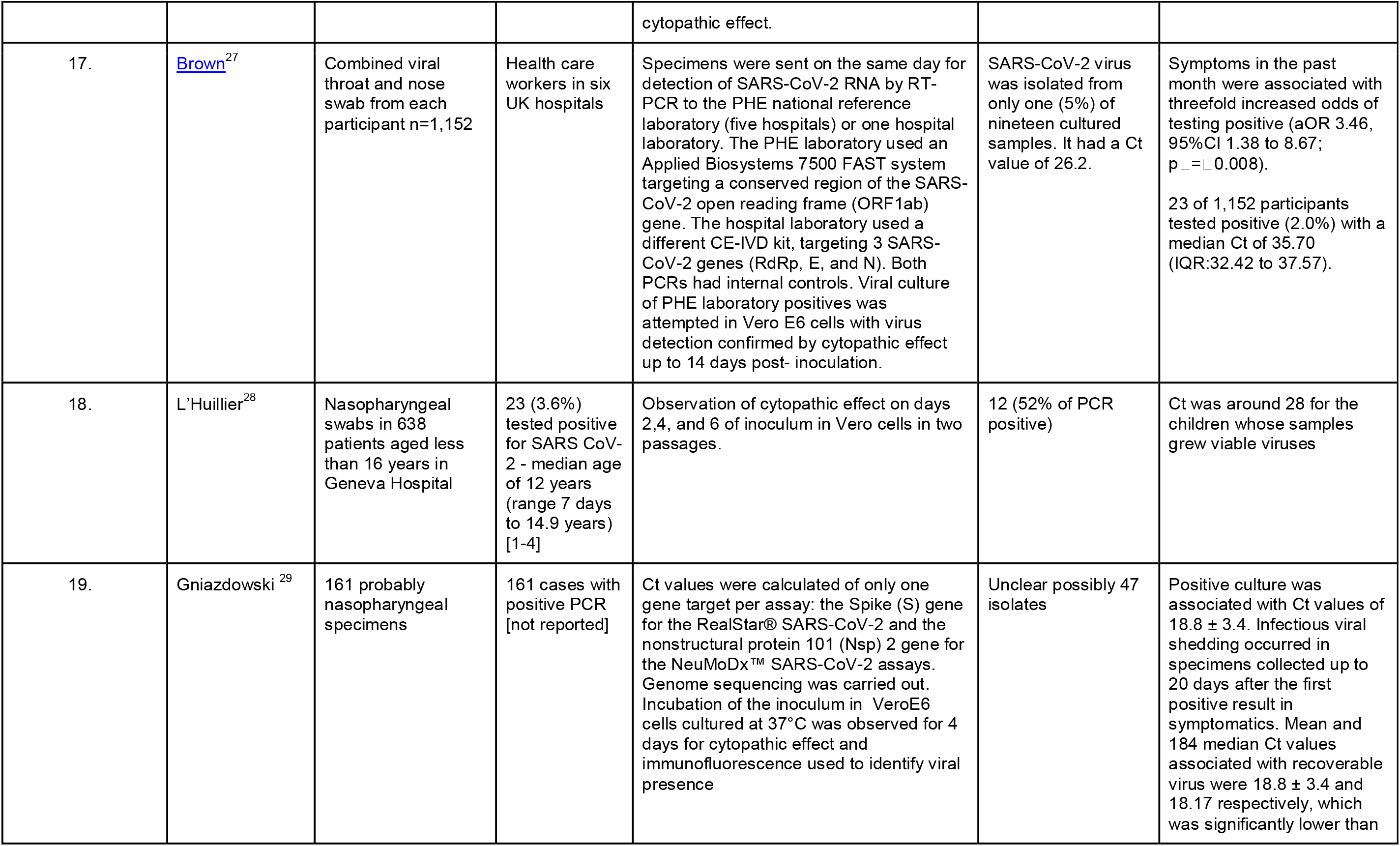
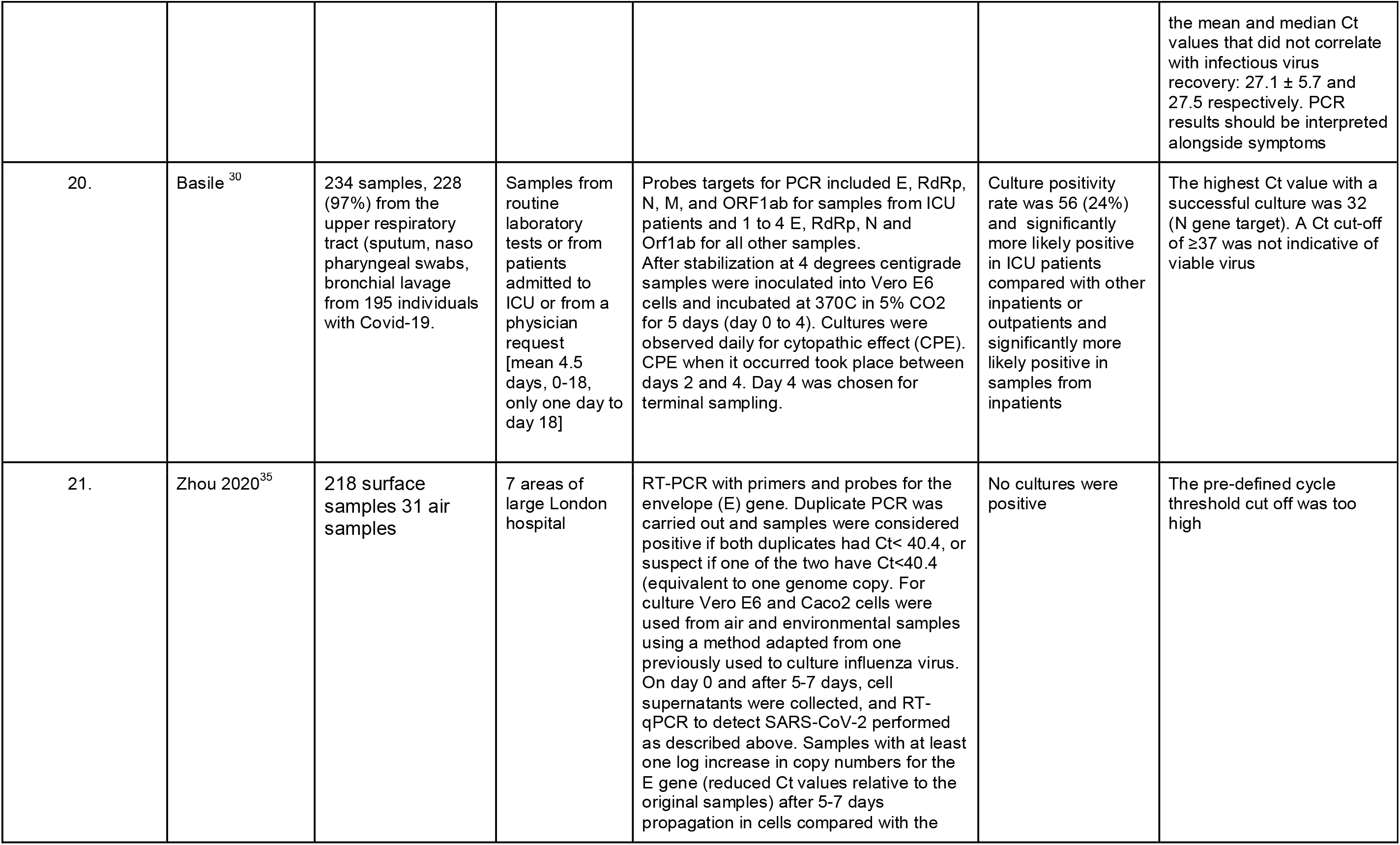
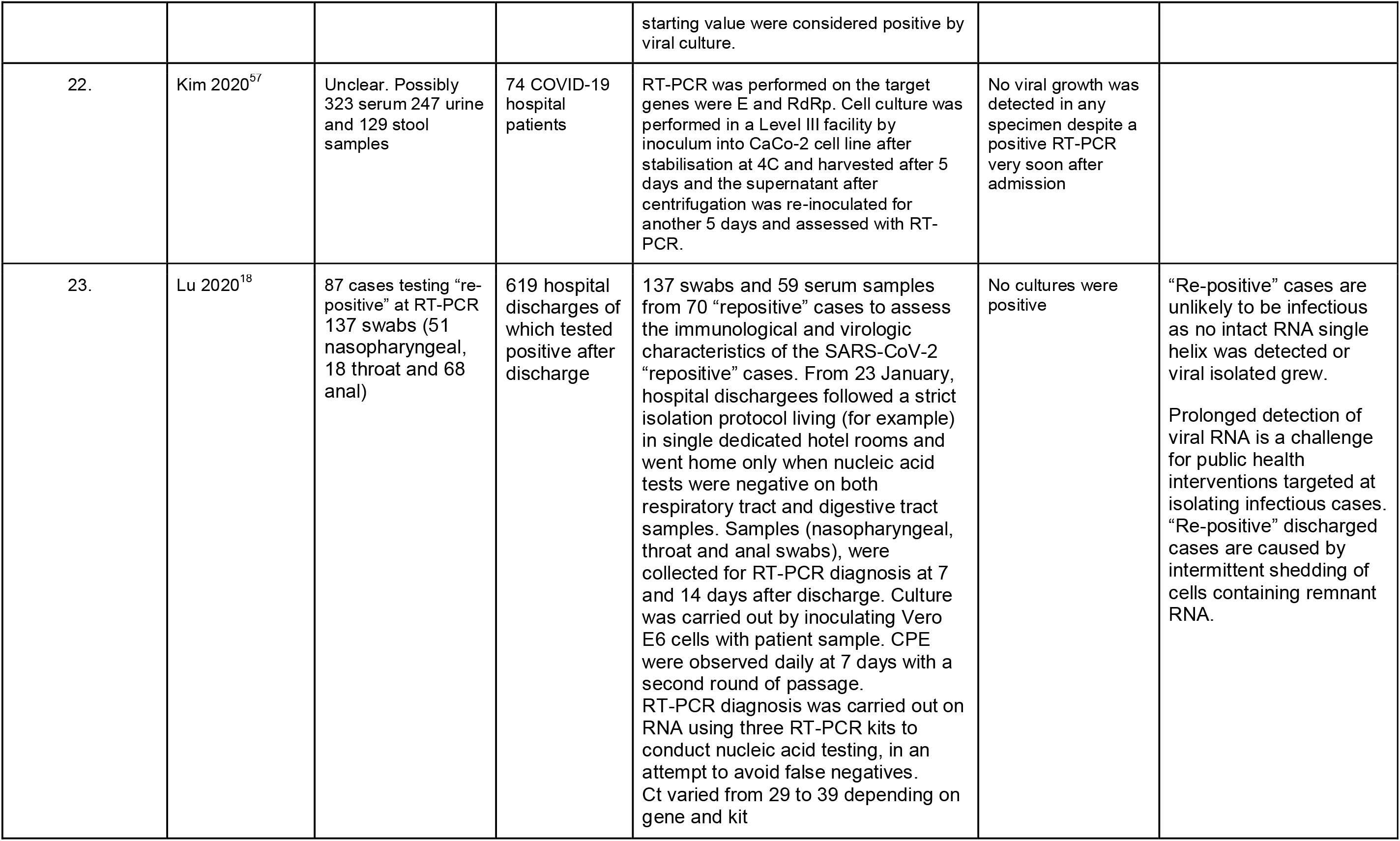
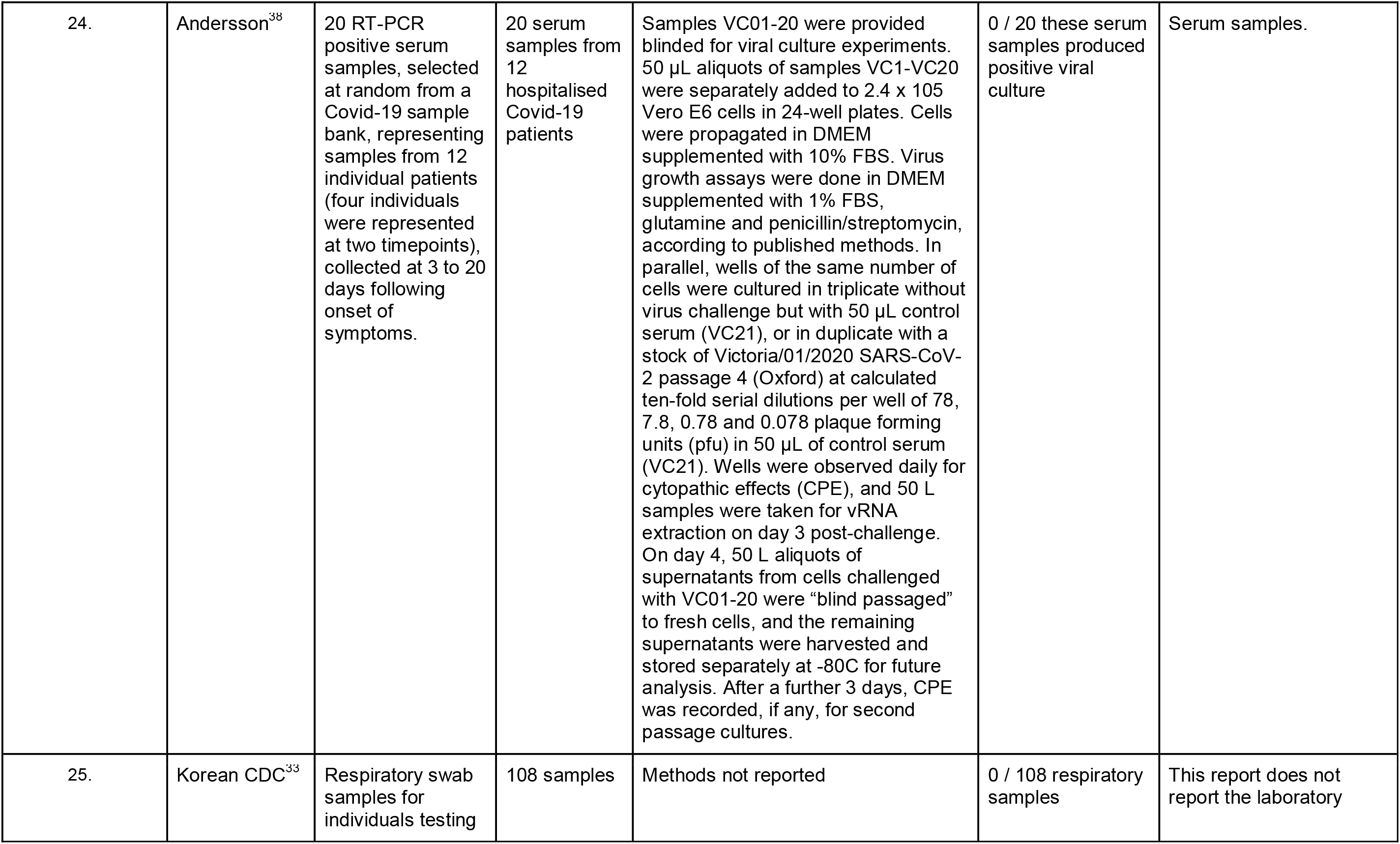
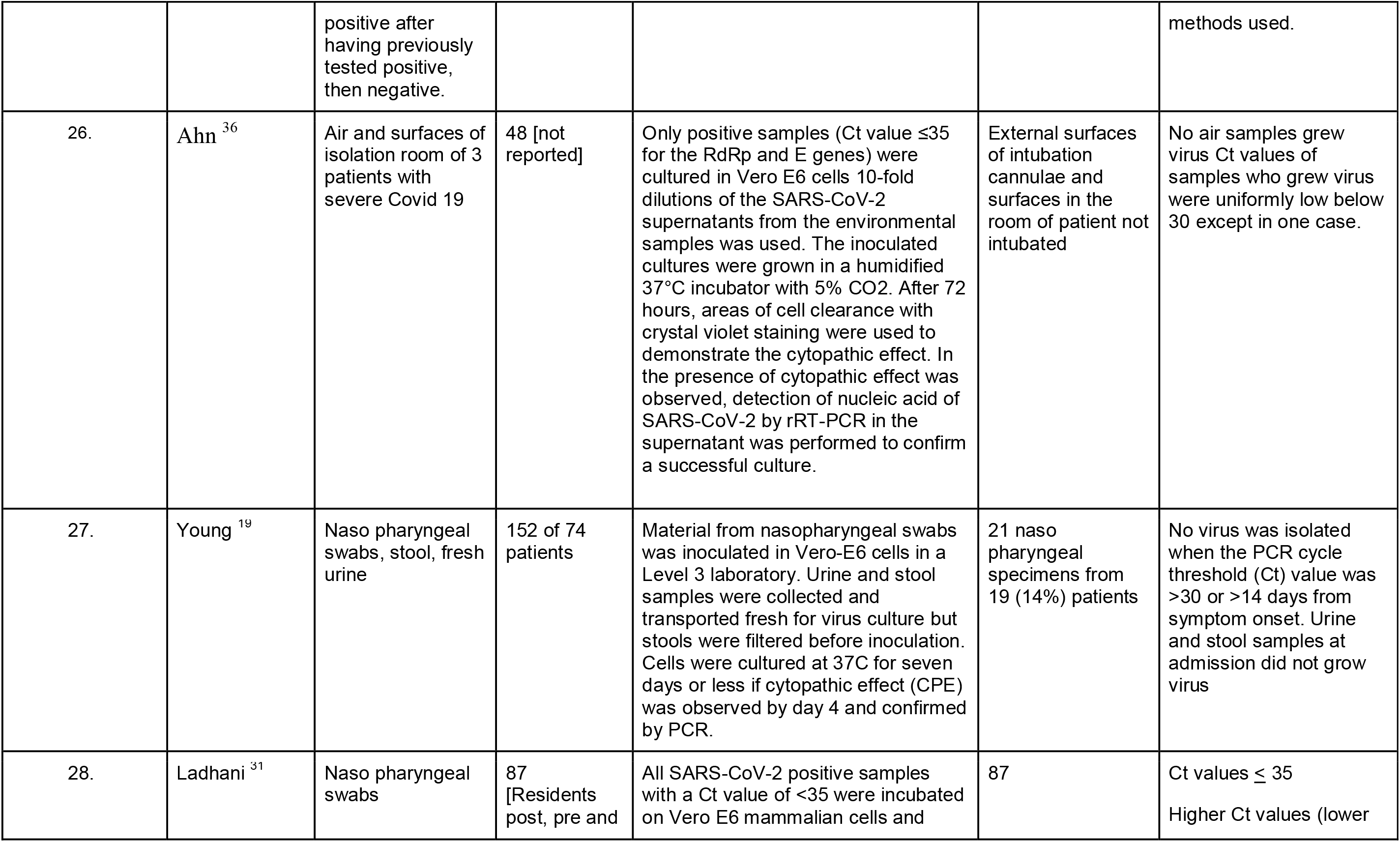
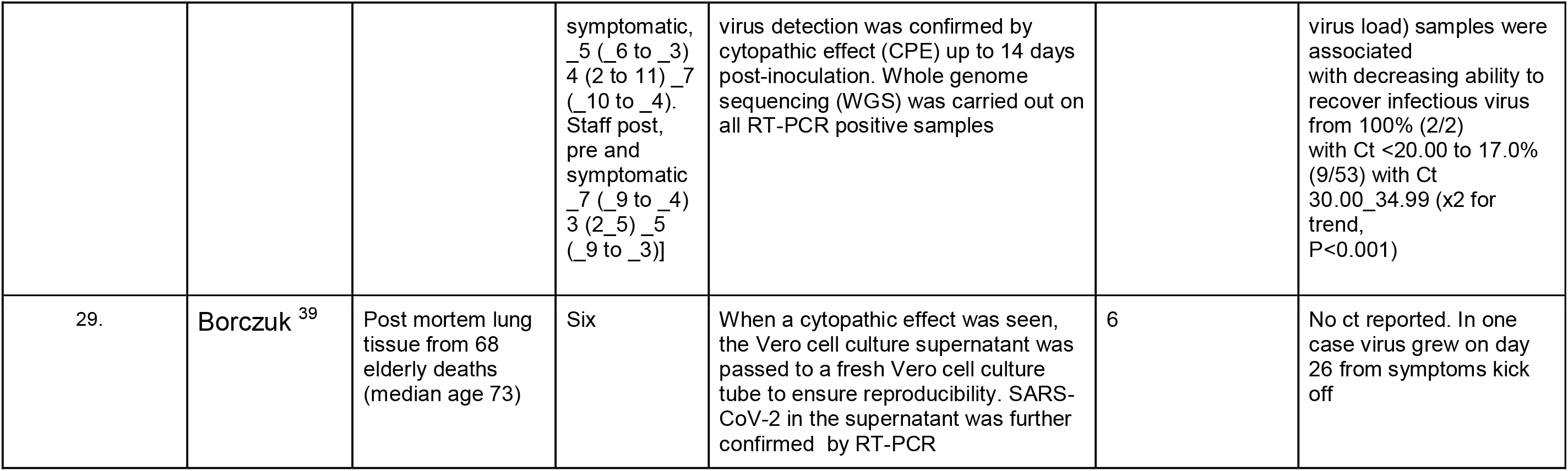
Characteristics of included studies. Key: STT = symptom onset to test date.

| Serial | Study | Samples (source) | Samples (n) [SST] | Culture methods | Culture Positive | Additional notes |
| --- | --- | --- | --- | --- | --- | --- |
| 1. | Bullard<sup>4</sup> | Nasopharyngeal (NP) or endotracheal (ETT) from COVID-19 patients (mean age 45 years) | 90 [0 to 7 days] | NP swabs and ETT specimens in viral transport media were stored at 4°C for 24-72 hours until they were tested for the presence of SARS-CoV-2 RNA using real-time RT-PCR targeting a 122nt portion of the Sarbecovirus envelope gene (E gene). Dilutions were placed onto the Vero cells in triplicate and incubated at 37°C with 5% CO2 for 96 hours. Following incubation of 4 days, cytopathic effect was evaluated under a microscope and recorded. | 26 | The range of symptoms onset to negative PCT was 21 days. Within this period, positive cultures were only observed up to day 8 post symptom onset |
| 2. | Huang<sup>5</sup> | Oropharyngeal (OP) or nasopharyngeal (NP) swabs, or sputum (SP) | 60 specimens from 50 cases [3,4 days mean but see table 1 for freeze thaw cycles delays] | SARS-CoV-2 cDNA was prepared using RNA extracted from the specimens of the first patient with confirmed COVID-19. RT was performed using the MMLV Reverse transcription kit. All procedures for viral culture were conducted in a biosafety level-3 facility. Vero-E6 and MK-2 (ATCC) cells were maintained in a virus culture medium and the cells were maintained in a 37°C incubator with daily observations of the cytopathic effect. | 12 OP, 9 NP and two from SP specimens were culturable | Specimens with high copy numbers of the viral genome, indicative of higher viral load, were more likely to be culturable. |
| 3. | Jeong<sup>11</sup> | Naso/oropharyngeal swabs, saliva, urine, and stool | 5 patients | Specimens positive by qPCR were subjected to virus isolation in Vero cells. Urine and stool samples were inoculated intranasally in ferrets and they evaluated the virus titers in nasal washes on 2, 4, 6, and 8 days post-infection (dpi). Immunofluorescence antibody assays were also done. | Naso/ oropharyngeal saliva, urine and stool<br>Samples were collected between days 8 to 30 of the clinical course. Viable SARS-CoV-2 was isolated from 1 naso / oropharyngeal swab. Ferrets inoculated with patient urine or stool were infected. SARS-CoV-2 was isolated from the nasal washes of the 2 urine-treated ferrets and one stool-treated ferret | Viral loads in urine, saliva, and stool samples were almost equal to or higher than those in naso / oropharyngeal swabs. After symptom resolution, patients shed viable virus in their saliva and urine up to day 15 of illness. |
| 4. | Qian<sup>20</sup> | Rectal tissue obtained from a surgical procedure was available. | 1 [1 to 3 days post op] | Ultrathin sections of tissue fixed in epoxy resin on formvar-coated copper grids were observed under electron microscope under 200kV. Immunohistochemical staining was used to establish expression and distribution of SARS-CoV-2 antigen. | 1 | No culture performed. Visualisation of virions in rectal tissue and detection of SARS-CoV-2 antigen in the rectal tissue. |
| 5. | Wang<sup>12</sup> | Bronchoalveolar fluid, sputum, feces, blood, and urine specimens from hospital in-patients with COVID-19 | 4 fecal samples with sufficiently high copy numbers from 1,070 specimens collected from 205 patients with COVID-19 (mean age of 44 years and 68% male [1 to 3 days from hospital admission]) | rRT-PCR targeting the open reading frame 1ab gene of SARS-CoV-2; cycle threshold values of rRT-PCR were used as indicators of the copy number of SARS-CoV-2 RNA in specimens with lower cycle threshold values corresponding to higher viral copy numbers. A cycle threshold value less than 40 was interpreted as positive for SARS-CoV-2 RNA. Four SARS-CoV-2 positive fecal specimens with high copy numbers were cultured, and then electron microscopy was performed to detect live virus. | 4 viewed by electron microscope | The details of how the 4 samples were cultured were not reported. The patients did not have diarrhoea. |
| 6. | Xiao F, Sun J<sup>13</sup> | Serial feces samples collected from 28 hospitalised COVID-19 patients: 3 samples from 3 RNA-positive patients were tested for possible viral | 3, one patient admitted day 7 post onset | Inoculation of Vero 6 cells. Cycle threshold values for the fecal sample were 23.34 for the open reading frame 1ab gene and 20.82 for the nucleoprotein gene. A cytopathic effect was visible in Vero E cells 2 days after a second-round passage. The researchers negatively stained culture supernatant and visualized | 2/3 (infectious virus was present in faeces from two cases) | Selection of samples is not entirely clear. |
|  |  | culture. |  | by transmission electron microscopy. Viral particles that were visible were spherical and had distinct surface spike protein projections, consistent with a previously published SARS-CoV2 image. |  |  |
| 7. | Arons <sup>23</sup> | nasopharyngeal and oropharyngeal swabs | 46 rRT-PCR–positive specimens [For asymptomatic median 4 days, Ct 23.1] | All rRT-PCR positive samples shipped to USA CDC for viral culture using Vero-CCL-81 cells. Cells showing cytopathic effects were used for SARS-CoV-2 rRT-PCR to confirm isolation and viral growth in culture. | 31 [no relation to symptoms presence. Culturable virus isolated from 6 days before to 9 days after symptom onset] |  |
| 8. | La Scola <sup>32</sup> | Naso pharyngeal swabs or sputum samples<br>Only Naso pharyngeal samples from the subsequent Jaafar et al letter. | 183 (4384 samples from 3466 patients) [not reported] | From 1,049 samples, 611 SARS-CoV-2 isolates were cultured. 183 samples testing positive by RT-PCR (9 sputum samples and 174 nasopharyngeal swabs) from 155 patients, were inoculated in cell cultures. SARS-CoV-2. RNA rtPCR targeted the E gene. Nasopharyngeal swab fluid or sputum sample were filtered and then inoculated in Vero E6 Cells. All samples were inoculated between 4 and 10 h after sampling and kept at + 4 °C before processing. After centrifugation they were incubated at 37 °C. They were observed daily for evidence of cytopathogenic effect. Two subcultures were performed weekly and scanned by electron microscope and then confirmed by specific RT-PCR targeting E gene. | Of the 183 samples inoculated in the studied period of time, 129 led to virus isolation. Of these 124 samples had detectable cytopathic effect between 24 and 96 h<br>The letter by Jaafar et al adds that 1941 SARS-Cov-2 30 isolate cultures were positive out 3 790 inoculated samples. These could be seen after the first inoculation or up to 2 blind subcultures. At at Ct of $\geq 34$ 2.6% of samples yielded a positive culture. | There was a significant relationship between Ct value and culture positivity rate: samples with Ct values of 13–17 all had positive culture. Culture positivity rate decreased progressively according to Ct values to 12% at 33 Ct. No culture was obtained from samples with Ct > 34. The 5 additional isolates obtained after blind subcultures had Ct between 27 and 34, thus consistent with low viable virus load. |
| 9. | Santarpia<sup>34</sup> | Windowsill and air, mean 7.3 samples per room. The percentage of PCR positive samples from each room was 40% -100% | 13 patients [days 5 to 9 and day 18 of isolation in a quarantine unit] | Vero E6 cells were used to culture virus from environmental samples. The cells were cultured in Dulbeccos's minimal essential medium (DMEM) supplemented with heat inactivated fetal bovine serum (10%), Penicillin/Streptomycin (10,000 IU/mL & 10,000 µg/mL) and Amphotericin B (25 µg/mL). | Possibly 2 with weak cytopathic effect | Isolates were from days 5 and 8 of occupancy of hospital/isolation rooms |
| 10. | Wölfel<sup>14</sup> | Saliva, nasal swabs, urine, blood and stool | 9 patients [2 to 4 days] | The average virus RNA load was $6.76 \times 10^5$ copies per the whole swab until day 5, and the maximum load was $7.11 \times 10^8$ copies per swab. The last swab sample that tested positive was taken on day 28 after the onset of symptoms. | Yes in respiratory samples, and indicative in stool | |
| 11. | Kujawski<sup>15</sup> (for The COVID-19 Investigation Team) | Nasopharyngeal (NP), oropharyngeal (OP), stool, serum and urine specimens | 9 from 9 patients | SARS-CoV-2 real-time PCR with reverse transcription (rRT-PCR) cycle threshold (Ct) values of virus isolated from the first tissue culture passage were 12.3 to 35.7 and for one patient, virus isolated from tissue culture passage 3 had a titer of $7.75 \times 10^6$ median tissue culture infectious dose per ml; these data were likely more reflective of growth in tissue culture than patient viral load. | 9 (including two non- hospitalised) | Viable SARS-CoV-2 was cultured at day 9 of illness (patient 10), but was not attempted on later specimens. SARS-CoV-2 rRT-PCR Ct values of virus isolated from the first tissue culture passage were 12.3 to 35.7. Mean Ct values in positive specimens were 17.0 to 39.0 for NP, 22.3 to 39.7 for OP and 24.1 to 39.4 for stool. All blood and urine isolates were negative. Ct values of upper respiratory tract specimens were lower in the first week of illness than the second in most patients, low Ct values continued into the second and third week of |

Viral cultures for COVID-19 infectivity assessment – a systematic review In: Analysis of the Transmission Dynamics of COVID-19: An Open Evidence Review
|  |  |  |  |  |  | illness. |
| --- | --- | --- | --- | --- | --- | --- |
| 12. | Zhang<sup>55</sup> | Stool | Unknown [not reported] | Vero cells were used for viral isolation from stool samples of Covid-19 patients. A 2019-nCoV strain was isolated from a stool specimen of a laboratory-confirmed COVID-19 severe pneumonia case, who experienced onset on January 16, 2020 and was sampled on February 1, 2020. The interval between sampling and onset was 15 days. The full-length genome sequence indicated that the virus had high-nucleotide similarity (99.98%) to that of the first isolated novel coronavirus isolated from Wuhan, China. In the Vero cells, viral particles with typical morphology of a coronavirus could be observed under the electron microscope. | 1 | We do not know what influenced successful virus culture e.g. methods optimal, or concentration of virus optimal. More information needed. |
| 13. | Xiao F. Tang M<sup>56</sup> | Esophageal, gastric, duodenal, and rectal tissues were obtained from 1 COVID-19 patients by endoscopy. | 1 plus an unknown additional number of fecal samples from RNA-positive patients. [not reported] | Histological staining (H&E) as well as viral receptor ACE2 and viral nucleocapsid staining were performed. | 1/1 RNA-positive patient. Positive staining of viral nucleocapsid protein was visualized in the cytoplasm of gastric, duodenal, and rectum glandular epithelial cell, but not in esophageal epithelium of the 1 patient providing these tissues. Additionally, positive staining of ACE2 and SARS-CoV-2 was also observed in gastrointestinal epithelium from other patients who tested positive for SARS-CoV-2 RNA in feces, results not shown. | Total sample numbers are not reported. |
| 14. | Yao<sup>16</sup> | Sputum (n=7), stool (n=3) and one nasopharyngeal sample | 11 patients admitted to hospital: 9 classified as serious or critical, 1 moderate, 1 mild symptoms [0 to 16 days] | The samples of the 11 patients involved in this study were collected during the early phase of the Covid-19 break out in China, dates ranging from 2nd of January to the 2nd of April 2020.<br>All except one of the patients had moderate or worse symptoms. Three patients had co-morbidities and one patient needed ICU treatment. Seven patients had sputum samples, one nasopharyngeal and three had stool samples<br>The samples were pre-processed by mixing with appropriate volume of MEM medium with 2% FBS, Amphotericin B, Penicillin G, Streptomycin and TPCK-trypsin. The supernatant was collected after centrifugation at 3000 rpm at room 434 temperature. Before infecting Vero-E6 cells, all collected supernatant was filtered using a 435 0.45 µm filter to remove cell debris etc.<br>Vero-E6 cells were infected with 11 viral isolates and quantitatively assessed their viral load at 1, 2, 4, 8, 24, and 48 hours post-infection (PI) and their viral cytopathic effects (CPE) at 48 and 72 hours PI and examined whether the viral isolates could successfully bind to Vero-E6 243 cells as expected. Super-deep sequencing of the 11 viral isolates on the Novaseq 6000 platform was performed. | 11 samples taken up to 16 days from admission to hospital. | Cultured viruses were inoculated in Vero cells. At 8 hours post-infection there was a significant decrease in Ct value (increases in viral load) for five isolates. At 24 hours significant decreases in the Ct values for all of the viral isolates were observed. Mutations of the viruses are also reported |
| 15. | Singanayagam<sup>25</sup> | 324 samples: nose, | 253 positive | Vero E6 cells were inoculated with | 133 (41%) samples | RT-PCR cycle threshold |
|  |  | throat, combined nose-and throat and nasopharyngeal swabs and aspirates | case [-10 to 60 days] | clinical specimens and incubated at 37 °C, 5% CO<sub>2</sub>. Cells were inspected for cytopathic effect daily up to 14 days. Presence of SARS-CoV-2 was confirmed by SARSCoV-2 nucleoprotein staining by enzyme immunoassay on infected cells. | (from 111 cases) | values correlate strongly with cultivable virus i.e. likelihood of infectiousness. Median Ct of all 324 samples was 31.15. Probability of culturing virus declines to 8% in samples with Ct > 35 and to 6% 10 days after onset and was similar in asymptomatic and symptomatic persons. Asymptomatic persons represent a source of transmissible virus but there is no difference in Ct values and culturability by age group. |
| 16. | Perera<sup>26</sup> | 68 specimens: nasopharyngeal aspirates combined with throat swab (n=49), nasopharyngeal aspirate (n=2), nasopharyngeal swab combined with throat swab (n=3), nasopharyngeal swab (n=2), sputum (n=11) and saliva (n=1). | 35 patients, 32 with mild disease [1 to 67 days] | Specimens were tested for sgRNA with $\geq 5$ log<sub>10</sub> N gene copies per mL. The complementary DNA obtained was subjected to PCR (40 cycles). Vero E6 cells were seeded and incubated for 24 hours in a CO<sub>2</sub> incubator. The culture medium was removed and 125 μL of the clinical specimen in virus transport medium diluted and was inoculated into 2 wells. After 2 hours incubation in a CO<sub>2</sub> incubator at 37°C, the plates were incubated at 37°C in a CO<sub>2</sub> incubator. A sample (100 μL) of supernatant was sampled for a quantitative real-time RT-PCR at 0 and 72 hours post inoculation. At 72 hours, cells were scraped into the supernatant and transferred onto fresh cells in 24-well plates and monitored for an additional 72 hours. A final quota of cells was collected for quantitative real-time RT-PCR. Cells were observed for cytopathic effect daily and harvested for testing if 25%–50% of cells showed a cytopathic effect. | 16/35 at a median 26 Ct | Culturable SARS CoV-2 and sub-genomic RNA (good indicator of replication) was rarely detectable beyond 8 days after onset of illness although virus RNA by RT-PCR remained for up to 70 days. |
| 17. | Brown<sup>27</sup> | Combined viral throat and nose swab from each participant n=1,152 | Health care workers in six UK hospitals | Specimens were sent on the same day for detection of SARS-CoV-2 RNA by RT-PCR to the PHE national reference laboratory (five hospitals) or one hospital laboratory. The PHE laboratory used an Applied Biosystems 7500 FAST system targeting a conserved region of the SARS-CoV-2 open reading frame (ORF1ab) gene. The hospital laboratory used a different CE-IVD kit, targeting 3 SARS-CoV-2 genes (RdRp, E, and N). Both PCRs had internal controls. Viral culture of PHE laboratory positives was attempted in Vero E6 cells with virus detection confirmed by cytopathic effect up to 14 days post- inoculation. | SARS-CoV-2 virus was isolated from only one (5%) of nineteen cultured samples. It had a Ct value of 26.2. | Symptoms in the past month were associated with threefold increased odds of testing positive (aOR 3.46, 95%CI 1.38 to 8.67; p<sub>L</sub>=0.008).<br>23 of 1,152 participants tested positive (2.0%) with a median Ct of 35.70 (IQR:32.42 to 37.57). |
| 18. | L'Huillier<sup>28</sup> | Nasopharyngeal swabs in 638 patients aged less than 16 years in Geneva Hospital | 23 (3.6%) tested positive for SARS CoV-2 - median age of 12 years (range 7 days to 14.9 years) [1-4] | Observation of cytopathic effect on days 2,4, and 6 of inoculum in Vero cells in two passages. | 12 (52% of PCR positive) | Ct was around 28 for the children whose samples grew viable viruses |
| 19. | Gniazdowski<sup>29</sup> | 161 probably nasopharyngeal specimens | 161 cases with positive PCR [not reported] | Ct values were calculated of only one gene target per assay: the Spike (S) gene for the RealStar® SARS-CoV-2 and the nonstructural protein 101 (Nsp) 2 gene for the NeuMoDx™ SARS-CoV-2 assays. Genome sequencing was carried out. Incubation of the inoculum in VeroE6 cells cultured at 37°C was observed for 4 days for cytopathic effect and immunofluorescence used to identify viral presence | Unclear possibly 47 isolates | Positive culture was associated with Ct values of 18.8 ± 3.4. Infectious viral shedding occurred in specimens collected up to 20 days after the first positive result in symptomatics. Mean and 184 median Ct values associated with recoverable virus were 18.8 ± 3.4 and 18.17 respectively, which was significantly lower than the mean and median Ct values that did not correlate with infectious virus recovery: $27.1 \pm 5.7$ and $27.5$ respectively. PCR results should be interpreted alongside symptoms |
| 20. | Basile <sup>30</sup> | 234 samples, 228 (97%) from the upper respiratory tract (sputum, naso pharyngeal swabs, bronchial lavage from 195 individuals with Covid-19. | Samples from routine laboratory tests or from patients admitted to ICU or from a physician request [mean 4.5 days, 0-18, only one day to day 18] | Probes targets for PCR included E, RdRp, N, M, and ORF1ab for samples from ICU patients and 1 to 4 E, RdRp, N and Orf1ab for all other samples. After stabilization at 4 degrees centigrade samples were inoculated into Vero E6 cells and incubated at 37°C in 5% CO<sub>2</sub> for 5 days (day 0 to 4). Cultures were observed daily for cytopathic effect (CPE). CPE when it occurred took place between days 2 and 4. Day 4 was chosen for terminal sampling. | Culture positivity rate was 56 (24%) and significantly more likely positive in ICU patients compared with other inpatients or outpatients and significantly more likely positive in samples from inpatients | The highest Ct value with a successful culture was 32 (N gene target). A Ct cut-off of $\geq 37$ was not indicative of viable virus |
| 21. | Zhou 2020<sup>35</sup> | 218 surface samples 31 air samples | 7 areas of large London hospital | RT-PCR with primers and probes for the envelope (E) gene. Duplicate PCR was carried out and samples were considered positive if both duplicates had Ct < 40.4, or suspect if one of the two have Ct < 40.4 (equivalent to one genome copy. For culture Vero E6 and Caco2 cells were used from air and environmental samples using a method adapted from one previously used to culture influenza virus. On day 0 and after 5-7 days, cell supernatants were collected, and RT-qPCR to detect SARS-CoV-2 performed as described above. Samples with at least one log increase in copy numbers for the E gene (reduced Ct values relative to the original samples) after 5-7 days propagation in cells compared with the starting value were considered positive by viral culture. | No cultures were positive | The pre-defined cycle threshold cut off was too high |
| 22. | Kim 2020<sup>57</sup> | Unclear. Possibly 323 serum 247 urine and 129 stool samples | 74 COVID-19 hospital patients | RT-PCR was performed on the target genes were E and RdRp. Cell culture was performed in a Level III facility by inoculum into CaCo-2 cell line after stabilisation at 4C and harvested after 5 days and the supernatant after centrifugation was re-inoculated for another 5 days and assessed with RT-PCR. | No viral growth was detected in any specimen despite a positive RT-PCR very soon after admission |  |
| 23. | Lu 2020<sup>18</sup> | 87 cases testing “re-positive” at RT-PCR 137 swabs (51 nasopharyngeal, 18 throat and 68 anal) | 619 hospital discharges of which tested positive after discharge | 137 swabs and 59 serum samples from 70 “repositive” cases to assess the immunological and virologic characteristics of the SARS-CoV-2 “repositive” cases. From 23 January, hospital discharges followed a strict isolation protocol living (for example) in single dedicated hotel rooms and went home only when nucleic acid tests were negative on both respiratory tract and digestive tract samples. Samples (nasopharyngeal, throat and anal swabs), were collected for RT-PCR diagnosis at 7 and 14 days after discharge. Culture was carried out by inoculating Vero E6 cells with patient sample. CPE were observed daily at 7 days with a second round of passage. RT-PCR diagnosis was carried out on RNA using three RT-PCR kits to conduct nucleic acid testing, in an attempt to avoid false negatives. Ct varied from 29 to 39 depending on gene and kit | No cultures were positive | “Re-positive” cases are unlikely to be infectious as no intact RNA single helix was detected or viral isolated grew.<br>Prolonged detection of viral RNA is a challenge for public health interventions targeted at isolating infectious cases. “Re-positive” discharged cases are caused by intermittent shedding of cells containing remnant RNA. |
| 24. | Andersson<sup>38</sup> | 20 RT-PCR positive serum samples, selected at random from a Covid-19 sample bank, representing samples from 12 individual patients (four individuals were represented at two timepoints), collected at 3 to 20 days following onset of symptoms. | 20 serum samples from 12 hospitalised Covid-19 patients | Samples VC01-20 were provided blinded for viral culture experiments. 50 µL aliquots of samples VC1-VC20 were separately added to 2.4 x 10<sup>5</sup> Vero E6 cells in 24-well plates. Cells were propagated in DMEM supplemented with 10% FBS. Virus growth assays were done in DMEM supplemented with 1% FBS, glutamine and penicillin/streptomycin, according to published methods. In parallel, wells of the same number of cells were cultured in triplicate without virus challenge but with 50 µL control serum (VC21), or in duplicate with a stock of Victoria/01/2020 SARS-CoV-2 passage 4 (Oxford) at calculated ten-fold serial dilutions per well of 78, 7.8, 0.78 and 0.078 plaque forming units (pfu) in 50 µL of control serum (VC21). Wells were observed daily for cytopathic effects (CPE), and 50 L samples were taken for vRNA extraction on day 3 post-challenge. On day 4, 50 L aliquots of supernatants from cells challenged with VC01-20 were “blind passaged” to fresh cells, and the remaining supernatants were harvested and stored separately at -80C for future analysis. After a further 3 days, CPE was recorded, if any, for second passage cultures. | 0 / 20 these serum samples produced positive viral culture | Serum samples. |
| 25. | Korean CDC<sup>33</sup> | Respiratory swab samples for individuals testing | 108 samples | Methods not reported | 0 / 108 respiratory samples | This report does not report the laboratory |

Viral cultures for COVID-19 infectivity assessment – a systematic review In: Analysis of the Transmission Dynamics of COVID-19: An Open Evidence Review
|  |  | positive after having previously tested positive, then negative. |  |  |  | methods used. |
| --- | --- | --- | --- | --- | --- | --- |
| 26. | Ahn <sup>36</sup> | Air and surfaces of isolation room of 3 patients with severe Covid 19 | 48 [not reported] | Only positive samples (Ct value $\leq 35$ for the RdRp and E genes) were cultured in Vero E6 cells 10-fold dilutions of the SARS-CoV-2 supernatants from the environmental samples was used. The inoculated cultures were grown in a humidified 37°C incubator with 5% CO<sub>2</sub>. After 72 hours, areas of cell clearance with crystal violet staining were used to demonstrate the cytopathic effect. In the presence of cytopathic effect was observed, detection of nucleic acid of SARS-CoV-2 by rRT-PCR in the supernatant was performed to confirm a successful culture. | External surfaces of intubation cannulae and surfaces in the room of patient not intubated | No air samples grew virus Ct values of samples who grew virus were uniformly low below 30 except in one case. |
| 27. | Young <sup>19</sup> | Naso pharyngeal swabs, stool, fresh urine | 152 of 74 patients | Material from nasopharyngeal swabs was inoculated in Vero-E6 cells in a Level 3 laboratory. Urine and stool samples were collected and transported fresh for virus culture but stools were filtered before inoculation. Cells were cultured at 37C for seven days or less if cytopathic effect (CPE) was observed by day 4 and confirmed by PCR. | 21 naso pharyngeal specimens from 19 (14%) patients | No virus was isolated when the PCR cycle threshold (Ct) value was >30 or >14 days from symptom onset. Urine and stool samples at admission did not grow virus |
| 28. | Ladhani <sup>31</sup> | Naso pharyngeal swabs | 87 [Residents post, pre and | All SARS-CoV-2 positive samples with a Ct value of <35 were incubated on Vero E6 mammalian cells and | 87 | Ct values $\leq 35$<br>Higher Ct values (lower |
|  |  |  | symptomatic, _5 (_6 to _3) 4 (2 to 11) _7 (_10 to _4). Staff post, pre and symptomatic _7 (_9 to _4) 3 (2_5) _5 (_9 to _3)] | virus detection was confirmed by cytopathic effect (CPE) up to 14 days post-inoculation. Whole genome sequencing (WGS) was carried out on all RT-PCR positive samples |  | virus load) samples were associated with decreasing ability to recover infectious virus from 100% (2/2) with Ct <20.00 to 17.0% (9/53) with Ct 30.00_34.99 (x2 for trend, P<0.001) |
| 29. | Borczuk<sup>39</sup> | Post mortem lung tissue from 68 elderly deaths (median age 73) | Six | When a cytopathic effect was seen, the Vero cell culture supernatant was passed to a fresh Vero cell culture tube to ensure reproducibility. SARS-CoV-2 in the supernatant was further confirmed by RT-PCR | 6 | No ct reported. In one case virus grew on day 26 from symptoms kick off |

All included studies were case series of **moderate quality** (Table 2. Quality of included studies). We could not identify a protocol for any of the studies. All the included studies had been either published or were available as preprints. All had been made public in 2020. We received five responses from authors regarding clarifying information (see Acknowledgments).

**Table 2.**
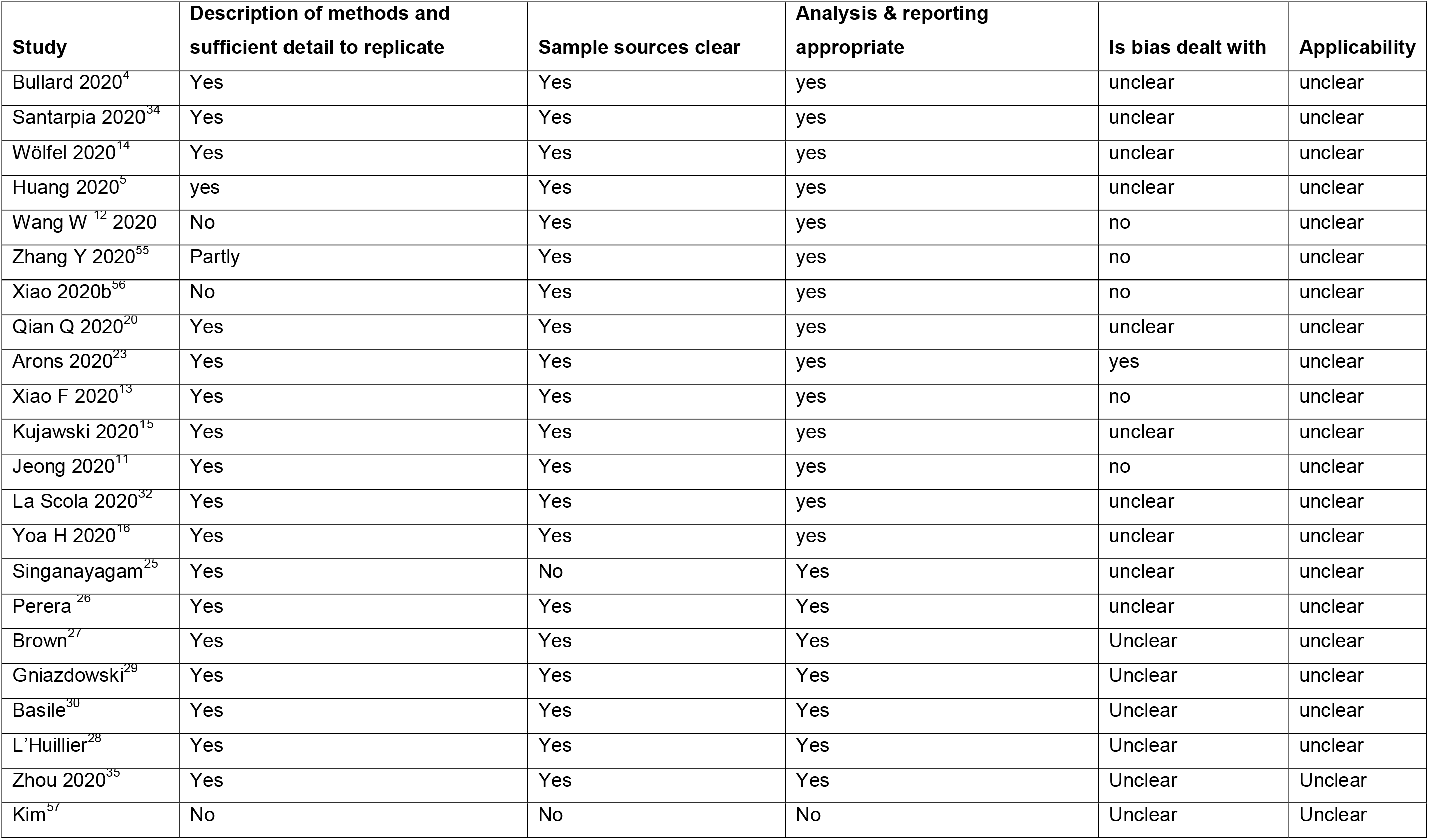
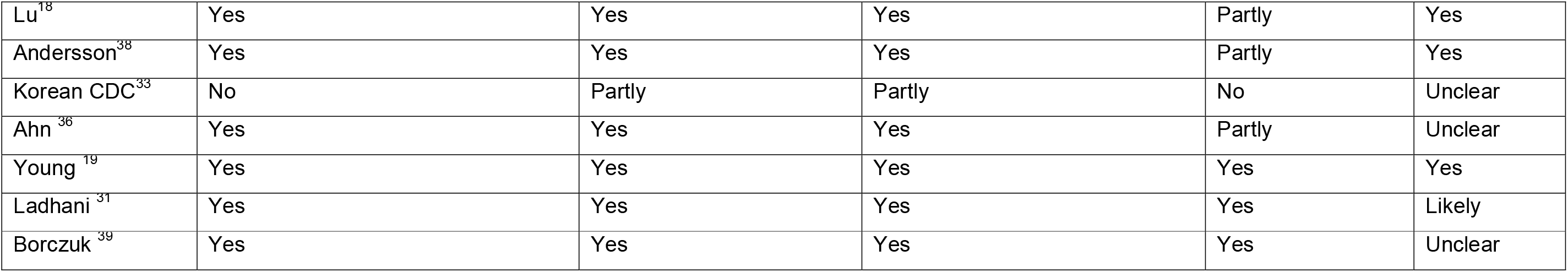
Quality of included studies

| **Study** | **Description of methods and sufficient detail to replicate** | **Sample sources clear** | **Analysis & reporting appropriate** | **Is bias dealt with** | **Applicability** |
| --- | --- | --- | --- | --- | --- |
| Bullard 2020<sup>4</sup> | Yes | Yes | yes | unclear | unclear |
| Santarpia 2020<sup>34</sup> | Yes | Yes | yes | unclear | unclear |
| Wölfel 2020<sup>14</sup> | Yes | Yes | yes | unclear | unclear |
| Huang 2020<sup>5</sup> | yes | Yes | yes | unclear | unclear |
| Wang W<sup>12</sup> 2020 | No | Yes | yes | no | unclear |
| Zhang Y 2020<sup>55</sup> | Partly | Yes | yes | no | unclear |
| Xiao 2020b<sup>56</sup> | No | Yes | yes | no | unclear |
| Qian Q 2020<sup>20</sup> | Yes | Yes | yes | unclear | unclear |
| Arons 2020<sup>23</sup> | Yes | Yes | yes | yes | unclear |
| Xiao F 2020<sup>13</sup> | Yes | Yes | yes | no | unclear |
| Kujawski 2020<sup>15</sup> | Yes | Yes | yes | unclear | unclear |
| Jeong 2020<sup>11</sup> | Yes | Yes | yes | no | unclear |
| La Scola 2020<sup>32</sup> | Yes | Yes | yes | unclear | unclear |
| Yoa H 2020<sup>16</sup> | Yes | Yes | yes | unclear | unclear |
| Singanayagam<sup>25</sup> | Yes | No | Yes | unclear | unclear |
| Perera<sup>26</sup> | Yes | Yes | Yes | unclear | unclear |
| Brown<sup>27</sup> | Yes | Yes | Yes | Unclear | unclear |
| Gniazdowski<sup>29</sup> | Yes | Yes | Yes | Unclear | unclear |
| Basile<sup>30</sup> | Yes | Yes | Yes | Unclear | unclear |
| L'Huillier<sup>28</sup> | Yes | Yes | Yes | Unclear | unclear |
| Zhou 2020<sup>35</sup> | Yes | Yes | Yes | Unclear | Unclear |
| Kim<sup>57</sup> | No | No | No | Unclear | Unclear |
| Lu<sup>18</sup> | Yes | Yes | Yes | Partly | Yes |
| Andersson<sup>38</sup> | Yes | Yes | Yes | Partly | Yes |
| Korean CDC<sup>33</sup> | No | Partly | Partly | No | Unclear |
| Ahn<sup>36</sup> | Yes | Yes | Yes | Partly | Unclear |
| Young<sup>19</sup> | Yes | Yes | Yes | Yes | Yes |
| Ladhani<sup>31</sup> | Yes | Yes | Yes | Yes | Likely |
| Borczuk<sup>39</sup> | Yes | Yes | Yes | Yes | Unclear |

### Studies using fecal samples

Nine studies assessed viral viability from fecal samples which were positive for SARS-CoV-2 based on RT-PCR result ^11-13 14-19^. One study reported infecting ferrets with stool supernatant^11^, two reported visual growth in tissue^12 20^ and five reported achieving viral replication^13-16^. One laboratory study^21^ found that SARS-CoV-2 infected human small intestinal organoids.

### Studies using respiratory samples

Sixteen studies on respiratory samples reported achieving viral isolation^4 22 11 23 24 14 15 16 25-28 19 29-31^. One study assessed 90 nasopharyngeal samples and cultured 26 of the samples, and positive cultures were only observed up to day eight post symptom onset; ^4^ another study obtained 31 cultures from 46 nasopharyngeal and oropharyngeal samples; ^23^ while 183 nasopharyngeal and sputum samples produced 124 cases in which a cytopathic effect was observed although the denominator of samples taken was unclear ^32^. Another study in health care workers in UK hospitals isolated one SARS Cov-2 from nineteen specimens in a situation of low viral circulation.^27^

Two more studies reported a clear correlation between symptoms onset, date of sampling, Ct and likelihood of viral culture. ^25 26^

L’Huillier and colleagues^28^ sampled nasopharyngeal swabs in 638 patients aged less than 16 years in a Geneva Hospital: 23 (3.6%) tested positive for SARS CoV-2 - median age of 12 years and 12 (52% were culture positive). The Ct was around 28 for the children whose samples grew viable viruses. Gniazdowski ^29^ probably assessed 161 nasopharyngeal specimens. A positive culture was associated with Ct values of 18.8 ± 3.4. Infectious viral shedding occurred in specimens (a Ct ≥ 23 yielded 8.5% of virus isolates).

Basile and colleagues ^30^ found a culture positivity rate of 24%, which was significantly more likely to be positive in ICU patients compared with other inpatients or outpatients.

A report by the Korean Centres for Disease Control failed to grow live viruses from 108 respiratory samples from “re-positives” i.e. people who had tested positive after previously testing negative^33^

Ladhani ^31^ and colleagues reported a successful culture rate of out 31 of 86 RT-PCR positive nasopharyngeal samples from six nursing home in London.

The largest number of positive culture came from the La Scola group publications^32^ with 1941 positive cultures from 3790 samples.

### Studies using environmental samples

Two possible positive cultures were obtained from 95 environmental samples in one study that assessed the aerosol and surface transmission potential of SARS-CoV-2 ^34^. Zhou and colleagues reported on samples taken from seven areas of a large London hospital. Despite apparent extensive air and surface contamination of the hospital environment, no infectious samples were grown^35^. For air samples, 2/31 (6.4%) were positive and 12/31 (39%) suspect for SARS-CoV-2 RNA but no virus was cultured. Similarly, 91 of 218 surface samples were suspect (42%) or 23 positive (11%) for SARS-CoV-2 RNA but no virus was cultured. The authors noted that a cut-off RT-PCR Ct > 30 was associated with non-infectious specimens.

Ahn and colleagues^36^ failed to grow live virus from an unspecified number of air samples in isolation rooms of patients with severe Covid-19 but were able to grow virus from swabs of hand rails, and the external surfaces of intubation cannulae.

### Mixed sources

Some of the studies labelled as mixed source samples are also reported in individual provenance breakdown in this text because of lack of clarity of the text.

Eight studies reported viral culture from mixed sources. Using 60 samples from 50 cases of Covid-19, viral culture was achieved from 12 oropharyngeal, nine nasopharyngeal and two sputum samples^5^. Jeong et al ^11^ who reported isolation live virus from a stool sample also reported that from of an unreported number of nasopharyngeal, oropharyngeal, saliva, sputum and stool samples, one viral culture was achieved: ferrets inoculated with these samples became infected; SARS-CoV-2 was isolated from the nasal washes of the two urine-treated ferrets and one stool-treated ferret^11^. An unreported number of samples from saliva, nasal swabs, urine, blood and stool collected from nine Covid-19 patients produced positive cultures and a possible specimen stool culture^14^. One study showed that from nine nasopharyngeal, oropharyngeal, stool, serum and urine samples, all nine were culturable, including two from non-hospitalised Covid-19 patients^15^.

Yao and colleagues cultured viable viral isolates from seven sputum samples, three stool samples and one nasopharyngeal sample of 11 patient aged 4 months to 71 years, indicating that the SARS-CoV-2 is capable of replicating in stool samples as well as sputum and the nasopharynx.^16^ All samples had been taken within 5 days of symptom onset. The authors also report a relationship between viral load (copy thresholds) and cytopathic effect observed in infected culture cells.^37^

Kim and colleagues reported no viral growth from and unclear number of serum, usirne and stool samples despite collection very soon after admission^17^. Lu and colleagues also reported no viral growth, however their specimens were from 87 cases tested “re-positive”.^18^

Young and colleagues ^19^ from Singapore had 21 positive cultures from 19 hospitalised patients in Singapore. No virus was isolated from samples with a Ct value >30, or when the sample was collected >14 days after symptoms onset. All positive cultures came from naso-pharyngeal samples, none of the 24 urine or 35 stool samples exhibited viral growth

### Blood cultures

In one study by Andersson^38^ et al 20 RT-PCR positive serum samples were selected at random from a Covid-19 sample bank, representing samples from 12 individual patients (four individuals were represented at two timepoints), collected at 3 to 20 days following onset of symptoms. None of the 20 serum samples produced a viral culture

### Post mortem study

One study on alveolar samples from 68 elderly deceased gre iable virus from 6 out 6 different samples, in one case on day 26 from symptom onset.^39^

### Duration of viral shedding

Nine studies report on the duration of viral shedding as assessed by PCR for SARS-CoV-2 RNA^4 11 20 13 14 15 13 25 40^. The minimum duration of RNA shedding detected by PCR was seven days reported in Bullard, the maximum duration of shedding was 35 days after symptom onset in Qian. Seven out of eight studies reported RNA shedding for longer than 14 days (see Table 3).

**Table 3.**
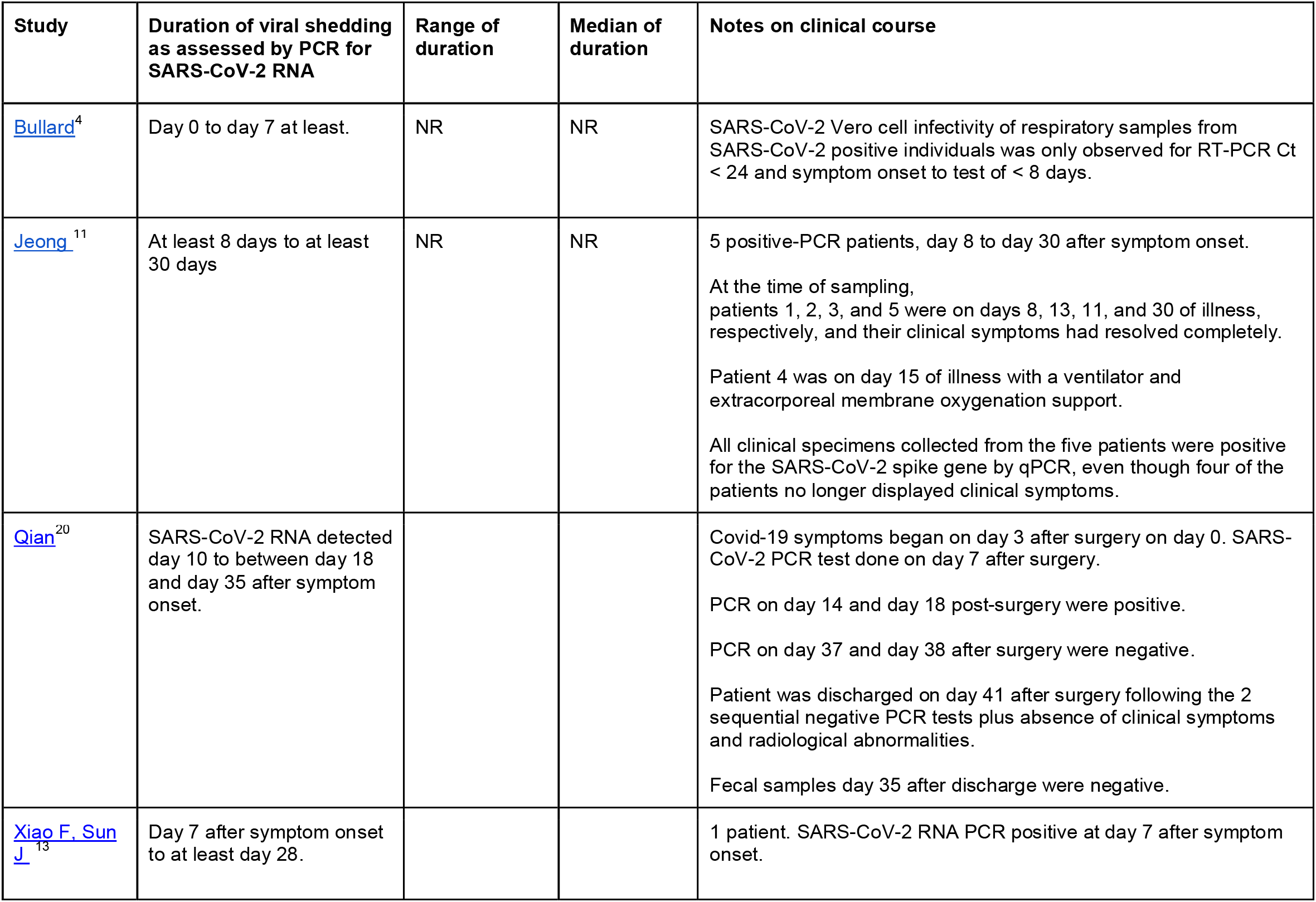
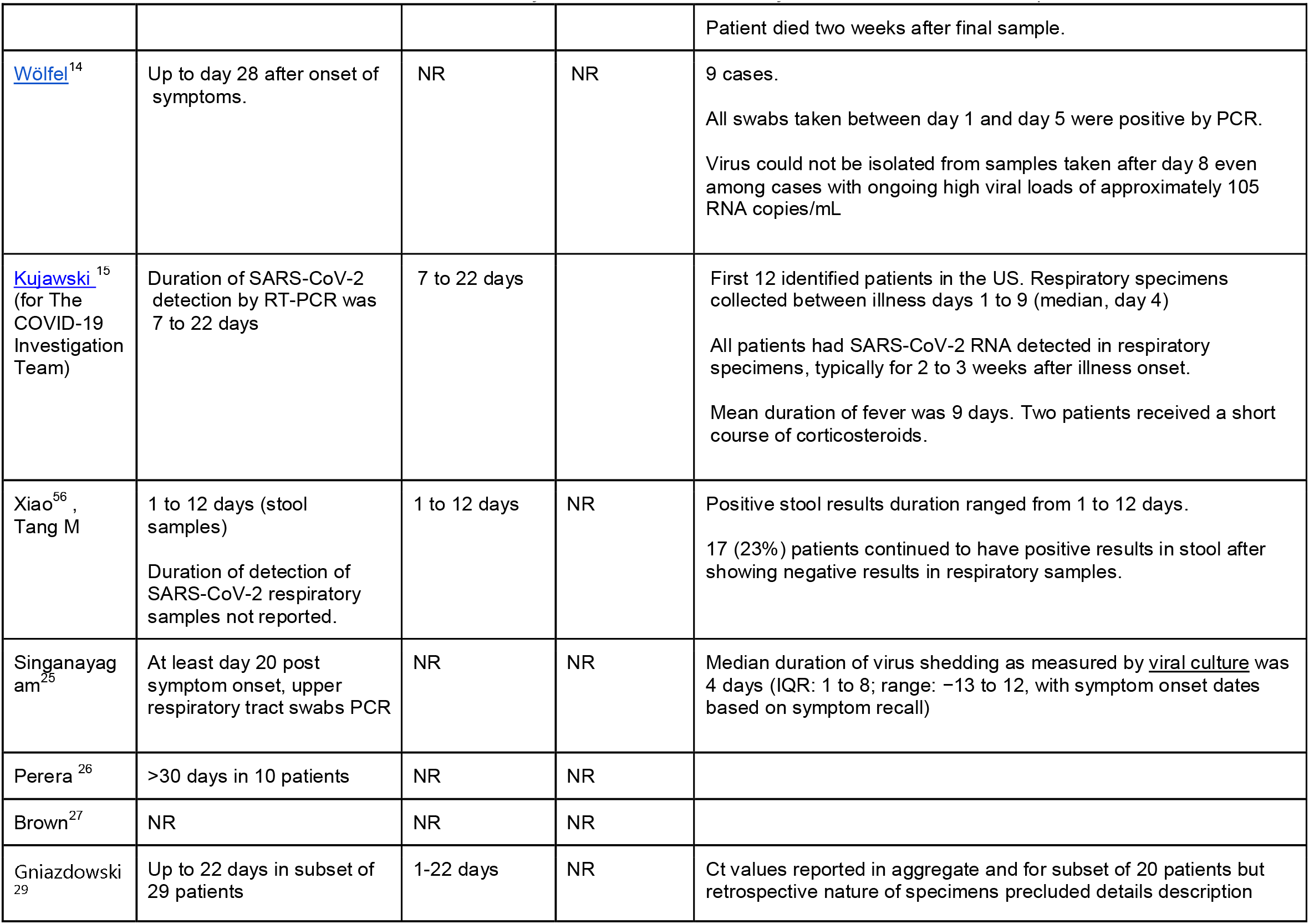
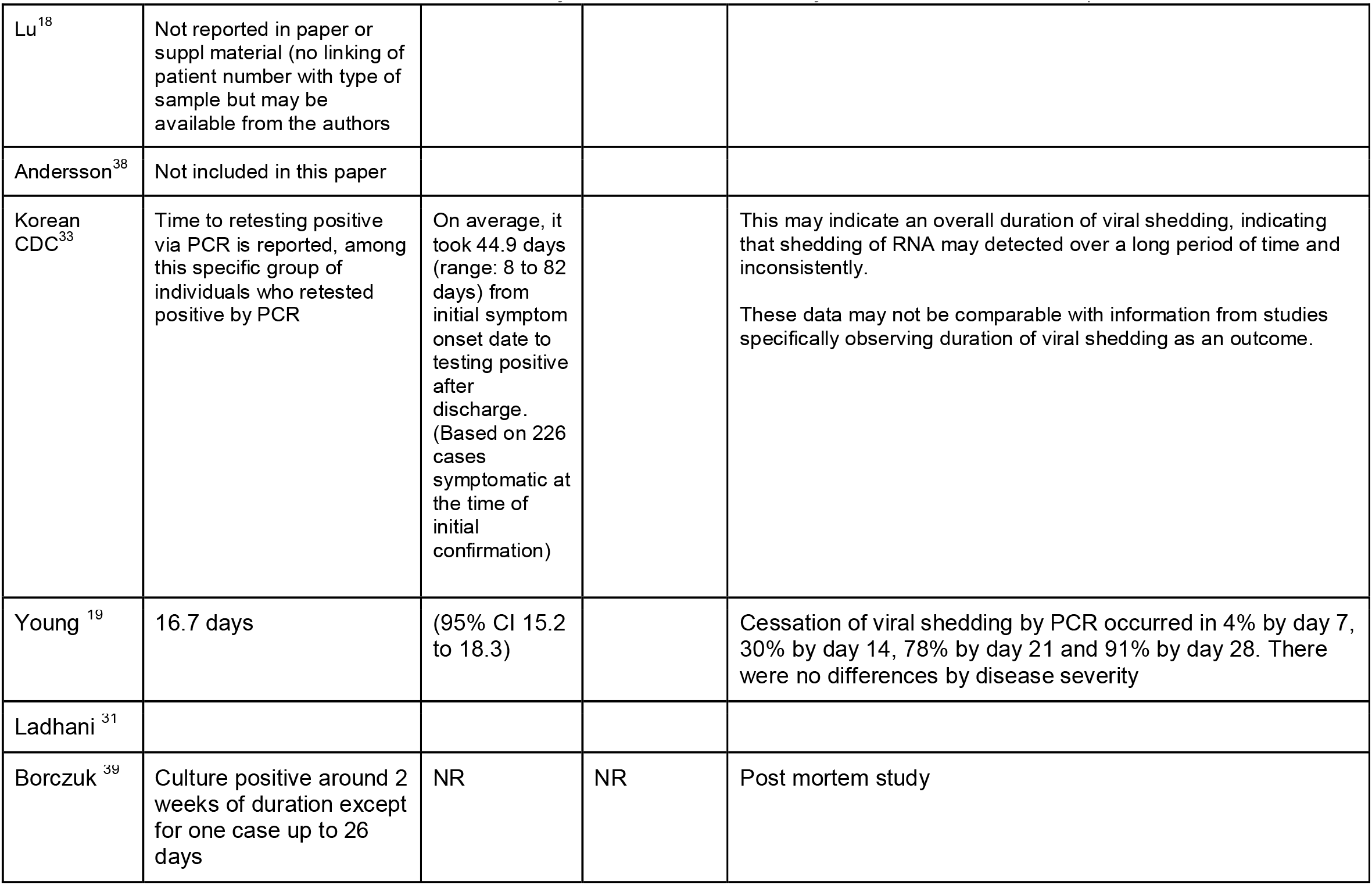
Duration of viral shedding in the included studies.

| Study | Duration of viral shedding as assessed by PCR for SARS-CoV-2 RNA | Range of duration | Median of duration | Notes on clinical course |
| --- | --- | --- | --- | --- |
| Bullard<sup>4</sup> | Day 0 to day 7 at least. | NR | NR | SARS-CoV-2 Vero cell infectivity of respiratory samples from SARS-CoV-2 positive individuals was only observed for RT-PCR Ct < 24 and symptom onset to test of < 8 days. |
| Jeong<sup>11</sup> | At least 8 days to at least 30 days | NR | NR | 5 positive-PCR patients, day 8 to day 30 after symptom onset.<br>At the time of sampling, patients 1, 2, 3, and 5 were on days 8, 13, 11, and 30 of illness, respectively, and their clinical symptoms had resolved completely.<br>Patient 4 was on day 15 of illness with a ventilator and extracorporeal membrane oxygenation support.<br>All clinical specimens collected from the five patients were positive for the SARS-CoV-2 spike gene by qPCR, even though four of the patients no longer displayed clinical symptoms. |
| Qian<sup>20</sup> | SARS-CoV-2 RNA detected day 10 to between day 18 and day 35 after symptom onset. |  |  | Covid-19 symptoms began on day 3 after surgery on day 0. SARS-CoV-2 PCR test done on day 7 after surgery.<br>PCR on day 14 and day 18 post-surgery were positive.<br>PCR on day 37 and day 38 after surgery were negative.<br>Patient was discharged on day 41 after surgery following the 2 sequential negative PCR tests plus absence of clinical symptoms and radiological abnormalities.<br>Fecal samples day 35 after discharge were negative. |
| Xiao F. Sun J<sup>13</sup> | Day 7 after symptom onset to at least day 28. |  |  | 1 patient. SARS-CoV-2 RNA PCR positive at day 7 after symptom onset. |
|  |  |  |  | Patient died two weeks after final sample. |
| Wölfel<sup>14</sup> | Up to day 28 after onset of symptoms. | NR | NR | 9 cases.<br>All swabs taken between day 1 and day 5 were positive by PCR.<br>Virus could not be isolated from samples taken after day 8 even among cases with ongoing high viral loads of approximately 105 RNA copies/mL |
| Kujawski<sup>15</sup><br>(for The COVID-19 Investigation Team) | Duration of SARS-CoV-2 detection by RT-PCR was 7 to 22 days | 7 to 22 days |  | First 12 identified patients in the US. Respiratory specimens collected between illness days 1 to 9 (median, day 4)<br>All patients had SARS-CoV-2 RNA detected in respiratory specimens, typically for 2 to 3 weeks after illness onset.<br>Mean duration of fever was 9 days. Two patients received a short course of corticosteroids. |
| Xiao<sup>56</sup>,<br>Tang M | 1 to 12 days (stool samples)<br>Duration of detection of SARS-CoV-2 respiratory samples not reported. | 1 to 12 days | NR | Positive stool results duration ranged from 1 to 12 days.<br>17 (23%) patients continued to have positive results in stool after showing negative results in respiratory samples. |
| Singanayagam<sup>25</sup> | At least day 20 post symptom onset, upper respiratory tract swabs PCR | NR | NR | Median duration of virus shedding as measured by <u>viral culture</u> was 4 days (IQR: 1 to 8; range: –13 to 12, with symptom onset dates based on symptom recall) |
| Perera<sup>26</sup> | >30 days in 10 patients | NR | NR |  |
| Brown<sup>27</sup> | NR | NR | NR |  |
| Gniazdowski<sup>29</sup> | Up to 22 days in subset of 29 patients | 1-22 days | NR | Ct values reported in aggregate and for subset of 20 patients but retrospective nature of specimens precluded details description |
| Lu<sup>18</sup> | Not reported in paper or suppl material (no linking of patient number with type of sample but may be available from the authors) |  |  |  |
| Andersson<sup>38</sup> | Not included in this paper |  |  |  |
| Korean CDC<sup>33</sup> | Time to retesting positive via PCR is reported, among this specific group of individuals who retested positive by PCR | On average, it took 44.9 days (range: 8 to 82 days) from initial symptom onset date to testing positive after discharge. (Based on 226 cases symptomatic at the time of initial confirmation) |  | This may indicate an overall duration of viral shedding, indicating that shedding of RNA may detected over a long period of time and inconsistently.<br>These data may not be comparable with information from studies specifically observing duration of viral shedding as an outcome. |
| Young<sup>19</sup> | 16.7 days | (95% CI 15.2 to 18.3) |  | Cessation of viral shedding by PCR occurred in 4% by day 7, 30% by day 14, 78% by day 21 and 91% by day 28. There were no differences by disease severity |
| Ladhani<sup>31</sup> |  |  |  |  |
| Borczuk<sup>39</sup> | Culture positive around 2 weeks of duration except for one case up to 26 days | NR | NR | Post mortem study |

Young et al^19^ reported that 91% of patients had ceased viral shedding by day 20 from symptom onset.

### Duration of live viral culture detection

The duration of live viral culture detection was much shorter than viral shedding. Wölfel et al ^14^reported that virus could not be isolated from samples taken after day 8 even among cases with ongoing high viral loads of approximately 105 RNA copies/mL.

Bullard et al similarly reported that SARS-CoV-2 Vero cell infectivity of respiratory samples from SARS-CoV-2 positive individuals was only observed for RT-PCR Ct < 24 and symptom onset to test of < 8 days^4^.

Singanayagam and colleagues ^25^ reported the median duration of virus shedding as measured by viral culture was 4 days (Inter Quartile Range: 1 to 8)^25^.

### The relationship between RT-PCR results and viral culture of SARS-CoV-2

Fifteen studies attempted to quantify the relationship between cycle threshold (Ct) and likelihood of culturing live virus^4 5 12 32 13 15 14 16 25 26 27 28-31^. Table 4 shows that nine studies analysed the relationship between Ct values and live viral culture^4 5 32 25 27 29 30 31 19^ and three quantified the mean log copies of detected virus and live culture^5 26 28^. All reported that Ct were significantly lower and log copies were significantly higher in those with live virus culture. Five studies reported no growth in samples based on a Ct cut-off value^4 5 27^.^19 31^ These values for no growth ranged from CT > 24^4^ to Ct ≥ 35^31^.

**Table 4:**
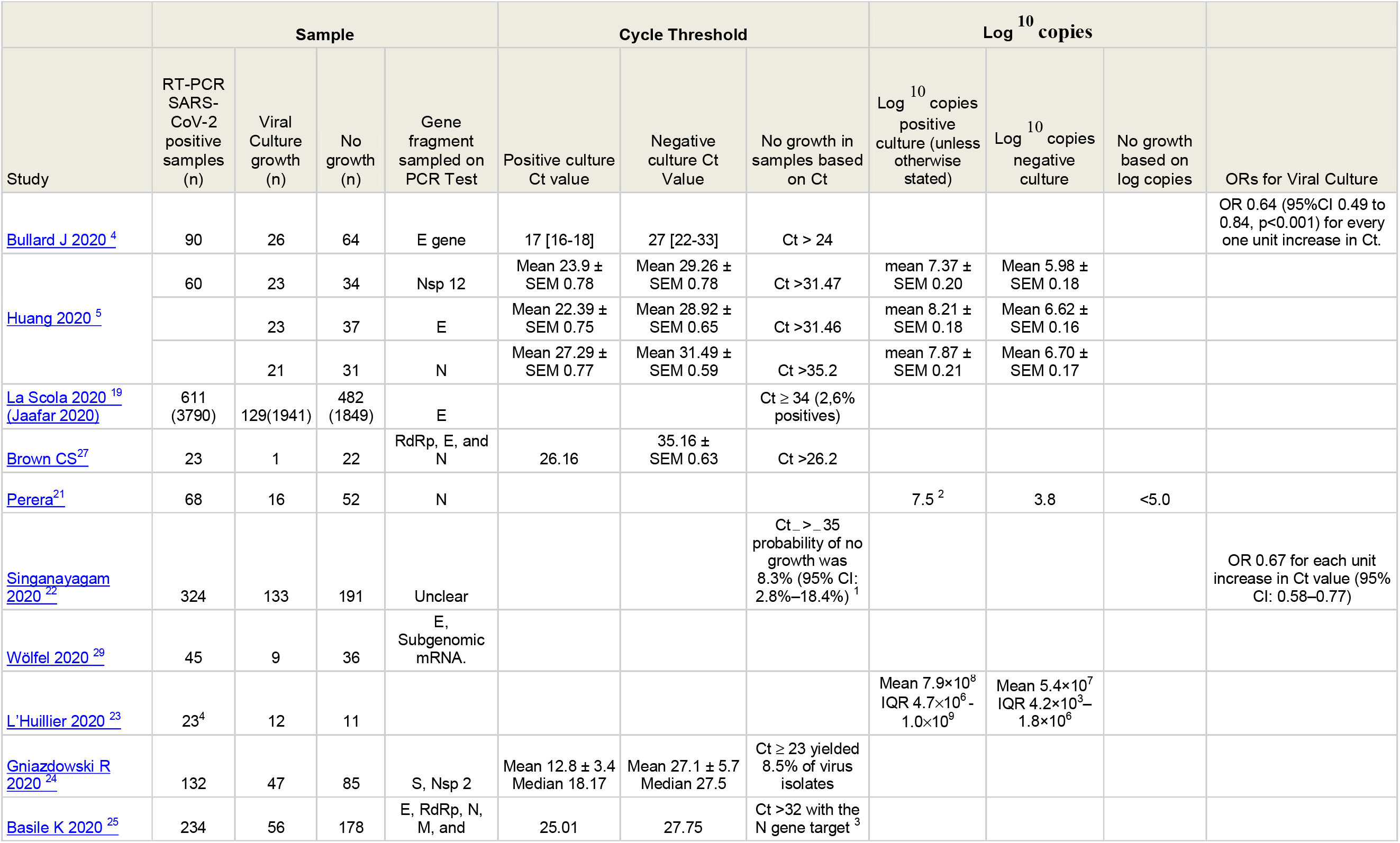
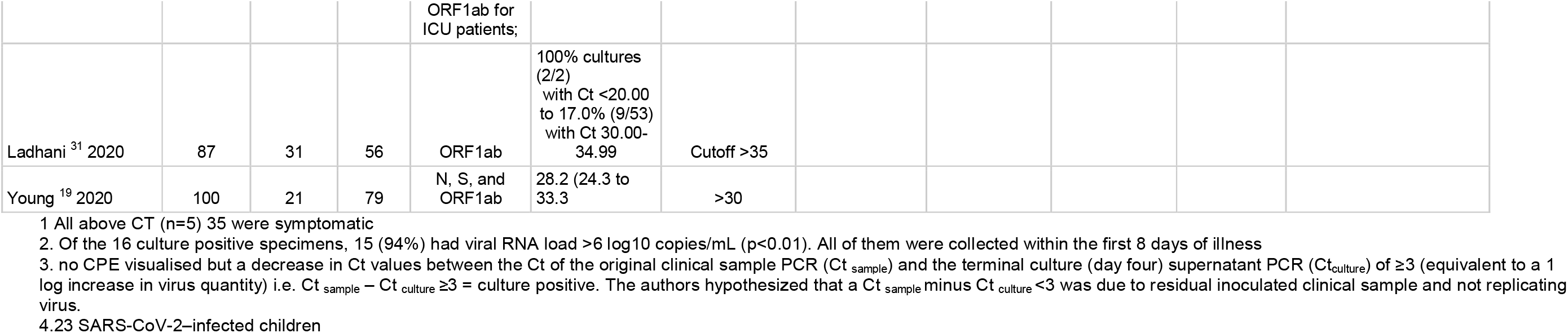
Relationship of PCR Cycle threshold and Log^10^ copies to Positive Viral Culture

| Study | Sample |  |  |  | Cycle Threshold |  |  | Log<sup>10</sup> copies |  |  | ORs for Viral Culture |
| --- | --- | --- | --- | --- | --- | --- | --- | --- | --- | --- | --- |
|  | RT-PCR SARS-CoV-2 positive samples (n) | Viral Culture growth (n) | No growth (n) | Gene fragment sampled on PCR Test | Positive culture Ct value | Negative culture Ct Value | No growth in samples based on Ct | Log<sup>10</sup> copies positive culture (unless otherwise stated) | Log<sup>10</sup> copies negative culture | No growth based on log copies |  |
| Bullard J 2020<sup>4</sup> | 90 | 26 | 64 | E gene | 17 [16-18] | 27 [22-33] | Ct > 24 |  |  |  | OR 0.64 (95%CI 0.49 to 0.84, p<0.001) for every one unit increase in Ct. |
| Huang 2020<sup>5</sup> | 60 | 23 | 34 | Nsp 12 | Mean 23.9 ± SEM 0.78 | Mean 29.26 ± SEM 0.78 | Ct >31.47 | mean 7.37 ± SEM 0.20 | Mean 5.98 ± SEM 0.18 |  |  |
|  |  | 23 | 37 | E | Mean 22.39 ± SEM 0.75 | Mean 28.92 ± SEM 0.65 | Ct >31.46 | mean 8.21 ± SEM 0.18 | Mean 6.62 ± SEM 0.16 |  |  |
|  |  | 21 | 31 | N | Mean 27.29 ± SEM 0.77 | Mean 31.49 ± SEM 0.59 | Ct >35.2 | mean 7.87 ± SEM 0.21 | Mean 6.70 ± SEM 0.17 |  |  |
| La Scola 2020<sup>19</sup><br>(Jaafar 2020) | 611 (3790) | 129(1941) | 482 (1849) | E |  |  | Ct ≥ 34 (2.6% positives) |  |  |  |  |
| Brown CS<sup>27</sup> | 23 | 1 | 22 | RdRp, E, and N | 26.16 | 35.16 ± SEM 0.63 | Ct >26.2 |  |  |  |  |
| Perera<sup>21</sup> | 68 | 16 | 52 | N |  |  |  | 7.5<sup>2</sup> | 3.8 | <5.0 |  |
| Singanavagam 2020<sup>22</sup> | 324 | 133 | 191 | Unclear |  |  | Ct > 35 probability of no growth was 8.3% (95% CI: 2.8%–18.4%)<sup>1</sup> |  |  |  | OR 0.67 for each unit increase in Ct value (95% CI: 0.58–0.77) |
| Wölfel 2020<sup>29</sup> | 45 | 9 | 36 | E, Subgenomic mRNA. |  |  |  |  |  |  |  |
| L'Huillier 2020<sup>23</sup> | 23<sup>4</sup> | 12 | 11 |  |  |  |  | Mean 7.9×10<sup>8</sup><br>IQR 4.7×10<sup>6</sup> - 1.0×10<sup>9</sup> | Mean 5.4×10<sup>7</sup><br>IQR 4.2×10<sup>3</sup> - 1.8×10<sup>6</sup> |  |  |
| Gniazdowski R 2020<sup>24</sup> | 132 | 47 | 85 | S, Nsp 2 | Mean 12.8 ± 3.4<br>Median 18.17 | Mean 27.1 ± 5.7<br>Median 27.5 | Ct ≥ 23 yielded 8.5% of virus isolates |  |  |  |  |
| Basile K 2020<sup>25</sup> | 234 | 56 | 178 | E, RdRp, N, M, and | 25.01 | 27.75 | Ct >32 with the N gene target<sup>3</sup> |  |  |  |  |

|  |  |  |  |  |  |  |
| --- | --- | --- | --- | --- | --- | --- |
|  |  |  |  | ORF1ab for ICU patients; |  |  |
| Ladhani <sup>31</sup> 2020 | 87 | 31 | 56 | ORF1ab | 100% cultures (2/2) with Ct <20.00 to 17.0% (9/53) with Ct 30.00-34.99 | Cutoff >35 |
| Young <sup>19</sup> 2020 | 100 | 21 | 79 | N, S, and ORF1ab | 28.2 (24.3 to 33.3 | >30 |

1 All above CT (n=5) 35 were symptomatic
2. Of the 16 culture positive specimens, 15 (94%) had viral RNA load >6 log<sub>10</sub> copies/mL (p<0.01). All of them were collected within the first 8 days of illness
3. no CPE visualised but a decrease in Ct values between the Ct of the original clinical sample PCR (Ct<sub>sample</sub>) and the terminal culture (day four) supernatant PCR (Ct<sub>culture</sub>) of ≥3 (equivalent to a 1 log increase in virus quantity) i.e. Ct<sub>sample</sub> – Ct<sub>culture</sub> ≥3 = culture positive. The authors hypothesized that a Ct<sub>sample</sub> minus Ct<sub>culture</sub> <3 was due to residual inoculated clinical sample and not replicating virus.
4.23 SARS-CoV-2–infected children

Singanayagam et al ^22^ reported the estimated probability of recovery of virus from samples with Ct < 35 was 8.3% (95% CI: 2.8%-18.4%). All donors above the Ct threshold of 35 (n=5) with live culture were symptomatic.

The study in London nursing homes by Ladhani and colleagues found no correlation between Ct values with presence or absence of symptoms in either residents or staff^31^, although nearly 50%of both categories were asymptomatic.

Huang and colleagues^5^ analysed the NSP, N and E gene fragments of the PCR result, which reported different cut-off thresholds depending on the gene fragment analysed^5^. No growth was found for the NSP 12 fragment at Ct > 31.47, whereas the value was higher for the N gene fragment at >35.2.

Bullard et al ^4^ reported a reduction in the odds ratio for culturing live virus of 0.64 for every one unit increase in Ct (95%CI 0.49 to 0.84, p<0.001). Similar to Bullard and colleagues, Singanayagam ^22^ reported a strong relationship between Ct value and ability to recover infectious virus: estimated OR of recovering infectious virus decreased by 0.67 for each unit increase in Ct value (95% CI: 0.58-0.77). This value is very close to that of other empirical studies (an increased Ct of 0.58 per day since symptoms started)^41^

Young et al^19^ reported no viral isolation from samples where the Ct value was >30, or when the sample was collected >14 days after symptoms onset.

## Discussion

Society is attempting to interrupt transmission of SARS-CoV-2 by identifying and isolating those who are sick and those who are infectious. As there are no Covid-19-specific mass treatments or preventive measures, such a strategy relies on our capability to identify infected and infectious persons with a reasonable amount of certainty to avoid isolation of those who pose little threat to the public health. An increasing body of evidence shows that such identification cannot be accurately achieved through the simplistic division of those who test positive and who do not, on the basis of the results of RT-PCR. The sensitivity and specificity of RT-PCR needs comparing to the gold standard of infectiousness: the capacity to grow live virus from a specimen.

Some of the authors of the studies in our review have attempted and successfully achieved culture of SARS-CoV-2 in the laboratory, using a range of respiratory, fecal or environmentally collected samples. However the simplistic dichotomous division into positive/negative is insufficient to accurately identify infectiousness as detection of viral RNA cannot support an inference of contagiousness^42^. The evidence shows that there is a positive relationship between lower cycle count threshold, likelihood of positive viral culture^43^ and date of symptom onset. Nowhere can this be seen as clearly as in the two studies assessing the infectiousness of “re-positives”, i.e. those COVID-19 cases who had been discharged from hospital after testing negative repeatedly and then testing positive after discharge: Lu 2020^18^, Korean CDC^33^.

In a very tightly designed and argued study Lu and colleagues tested four hypotheses for the origin of “re-positives”^18^. After discarding the first two (re-infection and latency) on the basis of their evidence, they reached the conclusions that the most plausible explanations were either contamination of the sample by extraneous material or identification in the sample of minute and irrelevant particles of SARS-CoV-2 debris representing virus long neutralised by the immune system.

Both explanations fit the facts, the others do not. It is very likely that a huge expansion in testing capability requires training protocols and precautions to avoid poor laboratory practice which are simply not possible in the restricted times of a pandemic. We equally know that weak positives (those with high Ct) are unlikely to be infectious, as a whole live virus is the prime requirement for transmission, not the fragments identified by PCR.

The purpose of viral testing is to assess the relation of the micro-organism and hazard to humans, i.e. its clinical impact on the individual providing the sample for primary care and the risk of transmission to others for public health. PCR on its own is unable to provide such answers. When interpreting the results of RT-PCR it is important to take into consideration the clinical picture, the cycle threshold value, the number of days from symptom onset to test (STT) and the specimen donor’s age ^44 42 43^. Several of our included studies assessed the relationship of these variables and there appears to be a time window during which shedding is at its highest with low cycle threshold and higher possibility of culturing a live virus, with viral load and probability of growing live virus of SARS-CoV2 peaking much sooner than that of SARS CoV-1 or MERS-CoV^42^. We propose that further work should be done on this with the aim of constructing a calibrating algorithm for PCR which are likely to detect infectious patients. PCR should be continuously calibrated against a reference culture in Vero cells in which cytopathic effect has been observed^4^. Confirmation of visual identification using methods, such as an immunofluorescence assay may also be relevant for some virus types^8^. Henderson and colleagues have called for a multicenter study of all currently manufactured SARS-CoV-2 nucleic acid amplification tests to correlate the cycle threshold values on each platform for patients who have positive and negative viral cultures. Calibration of assays could then be done to estimate virus viability from the cycle threshold with some certainty.^45^

Ascertainment of infectiousness is all the more important as there is good evidence of viral RNA persistence across a whole range of different viral RNA disease with little or no infectivity in the post infectious phase on MERS^46^, measles ^47^, other coronoviridae^48^, HCV and a variety of animal RNA viruses^48^. In one COVID-19 (former) case this persisted until day 78 from symptoms onset with a very high Ct ^41^ but no culture growth, showing its lack of infectiousness.

We are unsure whether SARS CoV-2 methods of cell culture have been standardised. Systems can vary depending upon the selection of the cell lines; the collection, transport, and handling of and the maintenance of viable and healthy inoculated cells^49^. We therefore recommend that standard methods for culture should be urgently developed and external quality assessment schemes be extended to to all laboratories offering testing for SARS CoV2.^50^. If identification of viral infectivity relies on visual inspection of cytopathogenic effect, then a reference culture of cells must also be developed to test recognition against infected cells. Viral culture may not be appropriate for routine daily results, but specialized laboratories should rely on their own ability to use viruses as controls, perform complete investigations when needed, and store representative clinical strains whenever possible^49^. In the absence of culture, ferret inoculation of specimen washings and antibody titres could also be used. It may be impossible to produce a universal Cycle threshold value as this may change with circumstances (e.g. hospital, community, cluster and symptom level), laboratory methods^51^ and the current evidence base is thin.

We suggest the WHO produce a protocol to standardise the use and interpretation of PCR and routine use of culture or animal model to continuously calibrate PCR testing, coordinated by designated Biosafety Level III laboratory facilities with inward directional airflow^52^. Further studies with standardised methods^51^ and reporting are needed to establish the magnitude and reliability of this association.

The results of our review are similar to those of the scoping review by Byrne and colleagues on infectivity periods^53^ and those of the living review by Cevick and colleagues^42^. Although the inclusion criteria are narrower than ours, the authors reviewed 79 studies on the dynamics, load and shedding for SARS CoV-1, MERS and SARS CoV-2 from symptoms onset. They conclude that although SARS-CoV-2 RNA shedding in respiratory (up to 83 days) and stool (35 days) can be prolonged, duration of viable virus is relatively shortlived (up to a maximum of 8 days from symptoms onset). Results that are consistent with Bullard et al who found no growth in samples with a cycle threshold greater than 24 or when symptom onset was greater than 8 days, and Wolfel et al^14^ who reported that virus could not be isolated from samples taken after day 8 even among cases with ongoing high viral loads.

The review by Rhee and colleagues also reaches conclusion similar to ours.^43^

The evidence is increasingly pointing to the probability of culturing live virus being related to the amount of viral RNA in the sample and, therefore, inversely related to the cycle threshold. Thus, blanket detection of viral RNA cannot be used to infer infectiousness. Length of excretion is also linked to age, male gender and possibly use of steroids and severity of illness.

The limits of our review are the low number of studies of relatively poor quality with lack of standardised reporting and lack of gold testing for each country involved in the pandemic. We plan to keep updating this review with emerging evidence.

## Conclusion

The current data are suggestive of a relation between the time from collection of a specimen to test, copy threshold, and symptom severity, but the quality of the studies limits drawing firm conclusions. We recommend that a uniform international standard for reporting of comparative SARS-CoV-2 culture with index test studies be produced. Particular attention should be paid to the relationship between the results of testing, clinical conditions and the characteristics of the source patients, description of flow of specimens and testing methods. Extensive training of operators and avoidance of contamination should take place on the basis of fixed and internationally recognised protocols. Defining cut off levels predictive of infectivity should be feasible and necessary for diagnosing viral respiratory infections using molecular tests^54^.

We will contact the corresponding authors of the 11 studies correlating Ct with likelihood of culture to assess whether it is possible to aggregate data and determine a firm correlation to aid decision making.

## Data Availability

All data included in the review are from publications or preprints. All extractions sheets with direct links to the source paper are available from https://www.cebm.net/evidence-synthesis/transmission-dynamics-of-covid-19/

## Acknowledgments

Drs Susan Amirian, Siyuan Ding, Long Rong and Sravanthi Parasato and Bernard La Scola provided additional information for this brief. Dr Maryanne DeMasi helped with reference identification.

## Disclaimer

The article has not been peer-reviewed. The views expressed in this commentary represent the views of the authors and not necessarily those of the host institution, the NHS, the NIHR, or the Department of Health and Social Care. The views are not a substitute for professional medical advice. It will be regularly updated see the evidence explorer at https://www.cebm.net/evidence-synthesis/transmission-dynamics-of-covid-19/ for regular updates to the evidence summaries and briefs.

## Authors

Tom Jefferson is a Senior Associate Tutor and Honorary Research Fellow, Centre for Evidence-Based Medicine, University of Oxford. Disclosure statement is here

Elizabeth Spencer is Epidemiology and Evidence Synthesis Researcher at the Centre for Evidence-Based Medicine. (Bio and disclosure statement here)

Jon Brassey is the Director of Trip Database Ltd, Lead for Knowledge Mobilisation at Public Health Wales (NHS) and an Associate Editor at the BMJ Evidence-Based Medicine.

Carl Heneghan is Professor of Evidence-Based Medicine, Director of the Centre for Evidence-Based Medicine and Director of Studies for the Evidence-Based Health Care Programme. (Full bio and disclosure statement here)

This work is licensed under a Creative Commons Attribution-Noncommercial 4.0 International License.

